# Short prescribed exercises can quantify upper limb functioning in neurodegenerative disease

**DOI:** 10.1101/2025.01.20.25320857

**Authors:** Marcin Straczkiewicz, Katherine M. Burke, Narghes Calcagno, Alan Premasiri, Kendall T. Carney, Fernando G. Vieira, Jukka-Pekka Onnela, James D. Berry

## Abstract

Digital health technologies (DHTs) can quantify movements in daily routines but rely heavily on participant adherence over prolonged wear times. Here we analyze accelerometry data from wrist-worn devices during short episodes of prescribed exercises (PEs) performed by 329 individuals living with amyotrophic lateral sclerosis (ALS) in a longitudinal study. We developed an automated and interpretable signal processing method to estimate upper limb movement counts, duration, intensity, and similarity of repetitions during PEs. Upper limb swing duration increased while intensity and similarity of movement decreased over time, indicating longer but less vigorous and less consistent upper limb movements over time. Intensity emerged as the most robust predictor of changes in upper limb function. The results suggest that PEs can effectively quantify upper limb function comparable to certain free-living measurements, providing greater flexibility in the clinical application of DHTs.

## 1. Introduction

Digital health technologies (DHTs), including smartphones and wearable devices, offer a transformative platform for frequent, remote health data collection. Recognized by the U.S. Department of Health and Human Services Food and Drug Administration (FDA)^1^, DHTs hold promise for improving trial efficiency and clinical decision-making by decentralizing data collection and reducing participant burden through at-home data capture.

DHTs can collect either passive data, from devices in free-living environments, or active data, from structured tasks such as questionnaires or specific activities. Both data types can contribute to clinical outcome assessments (COAs), which measure patient experiences, functional ability, or survival, and may serve as clinically meaningful endpoints in trials. DHTs have been successfully used across diverse populations and clinical settings^2,3^, to quantify physical activity and functional status at various stages of disease severity^4–13^. Additionally, early findings suggest that DHTs may also help aid in early detection of neurological diseases, potentially opening the door for neuroprotective interventions^14^.

Passive data collection with DHTs enables continuous, large-scale, and real-world monitoring with minimal participant burden^15,16^. However, participant adherence tends to decline over time^17,18^, and identifying patterns or accounting for non-wear periods can be challenging. On the other hand, in-clinic measurements rely on brief, intermittent tests to capture changes in function. While these tests have sound psychometric properties, they require periodic clinic visits. An intermediate approach could be to use DHTs to collect data during short, standardized movement regimens at home, thus minimizing participant burden and ensuring consistent, analyzable movements.

Amyotrophic lateral sclerosis (ALS) is a neurodegenerative disease that causes progressive muscle weakness, functional decline, impaired mobility, and difficulty with daily activities^19–21^. Historically, ALS research has relied on outcome measures including quantitative strength testing, vital capacity, and the ALS Functional Rating Scale-Revised (ALSFRS-R), a questionnaire administered by trained personnel. More recently, digital COAs have been used to quantify behavioral and functional changes over time in people with ALS^10^. Passive DHT data collection has typically focused on total activity volume, real-world gait assessment, and upper limb movements^12,22–28^, often requiring prolonged wear periods for continuous data acquisition.

In this study, we introduce a new approach to quantify upper limb function in ALS. We focus on brief, prescribed exercises (PEs) data collected using wrist-won accelerometer in a longitudinal study of community-dwelling participants with ALS. In our analysis, we determine the association between PEs metrics, time, and ALSFRS-R. By capturing data during both free-living and PEs, we compare movement metrics across the two settings. Finally, we test whether shorter exercise tasks are useful to quantify ALS disease progression.

## 2. Methods

### 2.1. Ethics declaration

This study adhered to the guidelines outlined in the Declaration of Helsinki – Ethical Principles for Medical Research Involving Human Subjects. The study protocol was approved by the Institutional Review Board (ADVARRA Center for IRB Intelligence (CIRBI)). All participants provided informed consent prior to any study procedures. There was no compensation for participation in the study.

### 2.2. Population and devices

The data used in this analysis were collected by ALS Therapy Development Institute as part of the ALS Research Collaborative Study, which up to date has enrolled over 600 individuals with ALS^29^. The program has assembled a comprehensive dataset that includes self-reported ALSFRS-R (ALSFRS-RSE) scores, digital physiologic data, speech recordings, accelerometer measurements, and a biorepository of skin biopsies, whole genome sequences, and plasma samples.

We analyzed data from 329 participants with ALS enrolled between September 2014 and January 2023, wearing ActiGraph GT3X+ accelerometers (ActiGraph, Pensacola, FL) on both their dominant and non-dominant wrists. These devices recorded continuous triaxial accelerometer measurements at a sampling frequency of 30 Hz with a dynamic range of ±6 g. In this decentralized study, devices were mailed to the participants, and they were instructed to wear the devices as much as possible for 7 days each month. During this time, they performed three PE sessions on alternating days consisting of five types of exercises each. The upper limb exercises involved shoulder abduction and adduction, raising the limb as high and as frequently as possible within a 45-second period, followed by a 45-second rest. A YouTube video^30^ was provided to guide participants through these PEs, which lasted approximately five minutes in total, including rest periods. Our analysis focused on the upper limb swings performed during these exercises.

### 2.3. Prescribed exercise quantification

#### 2.3.1. Limb swing detection

##### Estimation of fundamental frequency

The PEs can be described in terms of the total count of limb swings, their average duration, intensity, and similarity. We quantified these metrics from the time series wearable device data, segmented into individual limb swings.

Exercise timing was self-reported by study participants with one-second accuracy. To segment swings, we first transformed the raw tri-axial accelerometer data into univariate vector magnitude, which is robust to signal deviations and translations resulting from the non-stationary spatial orientation of the sensing device (**Figure 1a**). Based on self-annotated exercise intervals from participants, limb swing periods were identified using 1-second non-overlapping windows where the maximum normalized (z-score) signal amplitude exceeded 0.5 SD for at least 10 consecutive seconds (**Figure 1b**).

**Figure 1.**
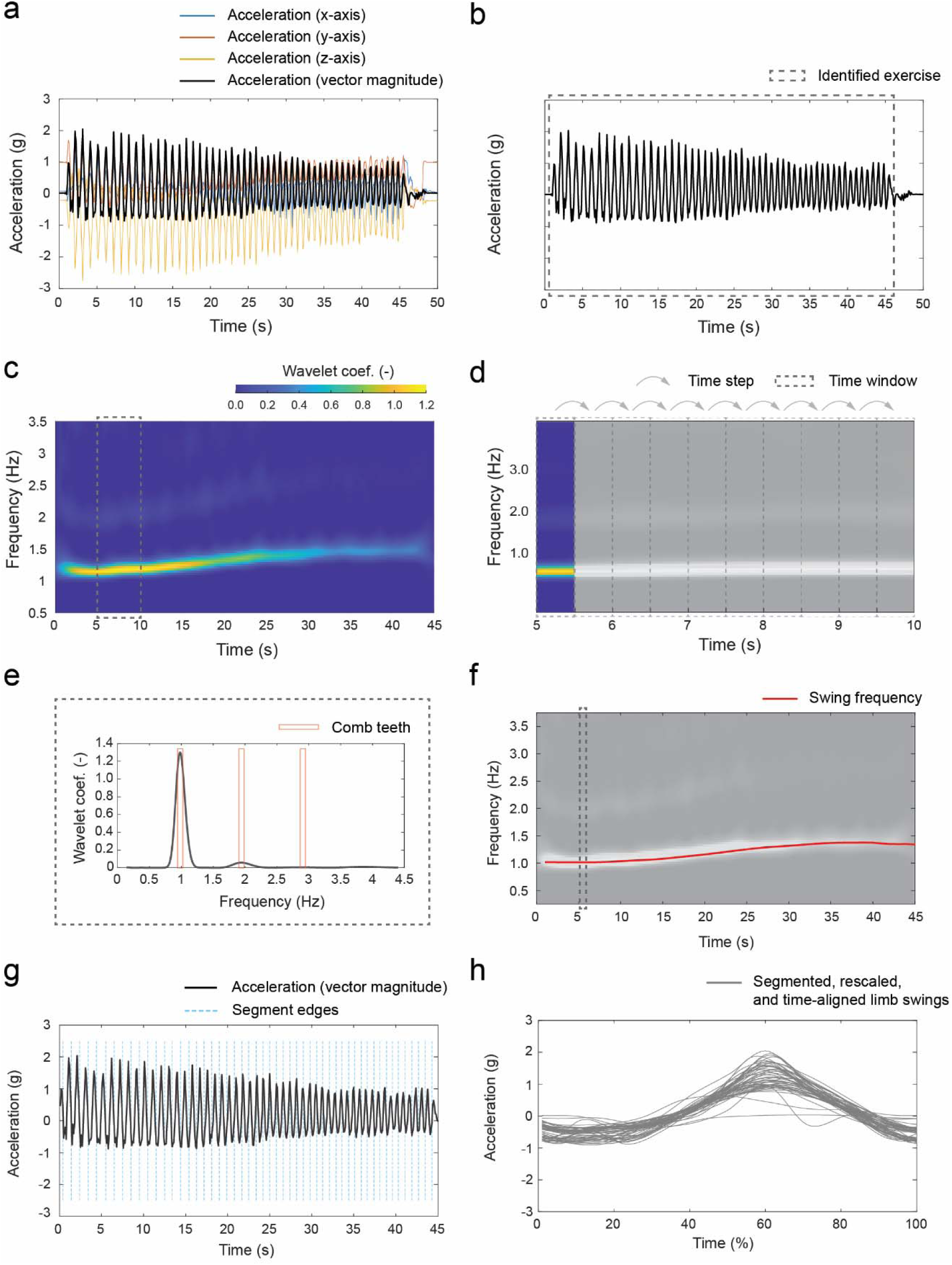
Workflow for quantifying limb swings from wearable device data. **a** Raw tri-axial accelerometer data were transformed into vector magnitude. **b** Limb swing periods were identified using 1-second windows with normalized signal amplitude above 0.5 SD. **c** Continuous wavelet transform (CWT) decomposed the signal into time-frequency components to capture fundamental swing frequency and harmonics. **d** Signal was split into non-overlapping windows. **e** Comb function screened all possible combinations of swing frequency and its higher harmonics. **f** Swing frequency was identified for each observation of the collected signal. **g** Swing frequency allowed creation of a sequence of segment edges. **h** Segmented signal fragments were rescaled and time aligned.

Next, we decomposed the signal into its time-frequency representation using continuous wavelet transform (CWT). This allowed us to uncover the dynamic characteristics of movement repetitions, with greater wavelet coefficients present at the fundamental frequency representing the limb swing frequency accompanied by its higher harmonics (i.e., peaks at multiples of the fundamental frequency) resulting from rapid changes in movement direction (**Figure 1c**). The time-frequency decomposition was then split into 0.5 s non-overlapping windows (**Figure 1d**), and the fundamental swing frequency was estimated from each window using a comb function (**Figure 1e**) as described previously for identifying stride frequency in gait and car vibrations while driving^31,32^.

##### Limb swing segmentation

We used the estimated fundamental frequency (**Figure 1f**) to segment limb swings. We performed this task by creating a sequence of observations corresponding to one full phase, such that each segment was bounded by adjacent observations and corresponded to an individual swing (**Figure 1g**). To align these segments and estimate their similarity, we first shrunk or stretched each segmented signal (we added a margin of approximately half of segment before and after the segment of interest) to match a length of arbitrary 100 observations and shifted it by a lag corresponding to the maximum cross-correlation between the interpolated segment and a sine wave (**Figure h**). To further improve this procedure, we identified the swing template with the highest mean cross-correlation with the remaining swing templates and repeated the alignment procedure using the identified template.

##### Outcome metrics

We calculated the total number of limb swings as the sum of all identified limb swing frequencies divided by two (correction for 0.5 s windows used to split the time-frequency decomposition)^33^ and the swing duration as the mean of the inverse of limb swing frequencies. Furthermore, we used segmented swing time series to calculate swing intensity and similarity. Specifically, we calculated the intensity as a mean acceleration of segmented and scaled limb swing time series, and we calculated the similarity as a mean pairwise correlation between all segmented, rescaled, and time-shifted limb swing time-series.

### 2.4. Free-living upper limb movements metrics

For comparison, we used a previously developed and validated approach for quantifying upper limb movement^24^. This approach focused on four aggregated upper limb movement metrics, *C_fe45_*, *C_sp45_*, *D_fe45_*, *D_sp45_*, which estimate the total number of forearm flexions and extensions by at least 45 degrees, the total number of forearm supinations and pronations by at least 45 degrees, the average duration of the ten fastest forearm flexions and extensions by at least 45 degrees, and the average duration of the ten fastest forearm supinations and pronations by at least 45 degrees, respectively.

### 2.5. Statistical analysis

The analytic dataset consisted of swing metrics and survey scores from participants that completed at least two exercise sessions on each limb and completed at least two corresponding ALSFRS-RSE surveys. The statistics on survey submission were computed for each participant, and we described them using mean, standard deviation (SD), median, and range.

The limb swing metrics were calculated for exercises with simultaneous wear of sensors on both wrist and were associated with the closest survey completed no more than 14 days from the exercise. If two surveys were equidistant from an exercise session, we used the earlier one.

In CWT, we used generalized Morse wavelet as the mother wavelet with symmetrical spread of coefficients in time and frequency domains^34^. The fundamental limb swing frequency was detected using a comb function with three teeth with a width of 0.1 Hz.

Pearson correlation was calculated using baseline observations, i.e., the first set of limb swing metrics and ALSFRS-RSE surveys. For surveys, we considered five scores: the sum of all responses to questions 1 through 12, denoted “Q1—12”, and a sum of responses for four areas of functioning: bulbar (“Q1—3”), fine motor (“Q4—6”), gross motor (“Q7—9”), and respiratory (“Q10—12”). The total ALSFRS-RSE is scored 0-48 with higher scores indicating higher level of function. Each of the four subdomains is scored 0-12, with higher values indicating higher level of function in that domain^35^. To determine the consistency of each metric during the first week of data, we calculated the intraclass correlation coefficient, ICC(3,1)^36^.

Further modelling considered measures and survey responses averaged on a weekly level. We used linear mixed-effect models (LMMs) with participant-specific slope and intercept to estimate baseline and weekly change of limb swing measures and ALSFRS-R scores. Additional LMMs were used to assess the association between measures and survey responses.

An alternative dataset used to compare the existing and new metrics was constructed from days with exercise data and with sensor wear time of at least 21 h. Sensor wear time was assessed using an established method proposed by Choi and colleagues^37^.

A sensitivity analysis was performed to determine whether measures from a prespecified number of limb swings were useful to characterize upper limb disease progression. In this scenario, metrics were quantified from 5, 10, 15, and 20 initial limb swing repetitions. Metrics were then fitted using additional LMMs, as described above.

For all analysis, the statistical significance level was set at 0.05. For the coefficient of determination, R-squared, we used grouped bootstrap with 1000 iterations to estimate its 95% confidence intervals.

### 2.6. Code availability

Data processing and statistical analyses were completed in MATLAB R2021a (MathWorks, Natick, MA). The code to estimate limb swing metrics is publicly available at (https://github.com/MStraczkiewicz/quantify_limb_swings). Code to estimate upper limb movement metrics used in comparative analysis has been published previously (https://github.com/MStraczkiewicz/quantify_forearm_movements).

## 3. Results

### 3.1. Population characteristics

Our analysis included 329 participants with ALS who contributed a total of 15467 epochs of accelerometer exercise data (about 200 hours) during at least two self-annotated upper limb exercise sessions (**Table 1**). The participants were predominantly right-handed (n=282, 85.71%) White (n=314, 95.44%) males (n=212, 64.44%). The average age of a study participants at enrollment was 54.20 (SD=11.01). On average, participants filled 31.81 (SD=26.64) surveys every 37.13 (20.06) days and completed 47.01 (47.09) exercises over 15.40 (15.65) weeks. Exercise duration was similar for non-dominant and dominant limbs, i.e., 46.49 (2.29) and 46.03 (2.11) s, respectively. The mean baseline total ALSFRS-RSE score was 40.32 (5.31), while mean baseline Q4—6 score (representing fine motor function) was 9.50 (2.34).

**Table 1.** Summary on population characteristics, collected surveys, and exercise statistics.

|  |  |
| --- | --- |
| Population characteristics |  |
| Total number of participants, N | 329 |
| Demographics |  |
| Age, mean (SD), median [min, max] | 54.20 (11.01) |
| Males, N (%) | 212 (64.44) |
| Race, N (%) |  |
| White | 314 (95.44) |
| Hispanic | 5 (1.52) |
| Asian | 5 (1.52) |
| Black | 2 (0.61) |
| Other | 3 (0.91) |
| Handedness, N (%) |  |
| Right | 282 (85.71) |
| Left | 46 (13.98) |
| Either | 1 (0.30) |
| **ALSFRS-RSE**, mean (SD) |  |
| Number of surveys, count | 31.81 (26.64) |
| Time between surveys, days | 37.13 (20.06) |
| **Exercise**, mean (SD) |  |
| Weeks with exercise per participant, count | 16.71 (16.15) |
| Time between weeks with exercises, weeks | 6.27 (2.84) |
| Annotated exercise per participant, count | 47.01 (47.09) |
| Exercise duration, non-dominant wrist, seconds | 46.49 (2.29) |
| Exercise duration, dominant wrist, seconds | 46.03 (2.11) |

### 3.2. Baseline associations

During the first week of observation, 259 (78.72 %) participants completed three non-dominant limb exercises, 46 (13.98 %) completed two, and 10 (3.04 %) completed one. Fourteen participants (4.26 %) did not perform any upper limb exercises. The intraclass correlation coefficients, ICC(3,1), between metrics derived from participants who completed three exercises indicated good to excellent consistency, with values of 0.819 for total limb swing count, 0.799 for duration, 0.955 for intensity, and 0.890 for similarity.

For the dominant wrist, the ICC(3,1) values were higher for count (0.861), duration (0.840), and intensity (0.946), but not for similarity (0.808). A similar distribution of exercise completion was observed: 257 participants (78.12%) completed three dominant limb exercises, 48 (14.59%) completed two, 10 (3.04%) completed one, and 14 (4.26%) performed none.

Correlation between three of the four swing metrics (count, duration, similarity) of the non-dominant limb and ALSFRS-RSE responses was very weak to weak, while the correlation between intensity and total score (Q1—12) and the fine motor functioning subdomain (Q4—6) was moderate, with values of 0.43 and 0.45, respectively (**Figure 2**), suggesting substantial between-patient heterogeneity. Similar correlation coefficients were computed for the dominant upper limb swings.

**Figure 2.**
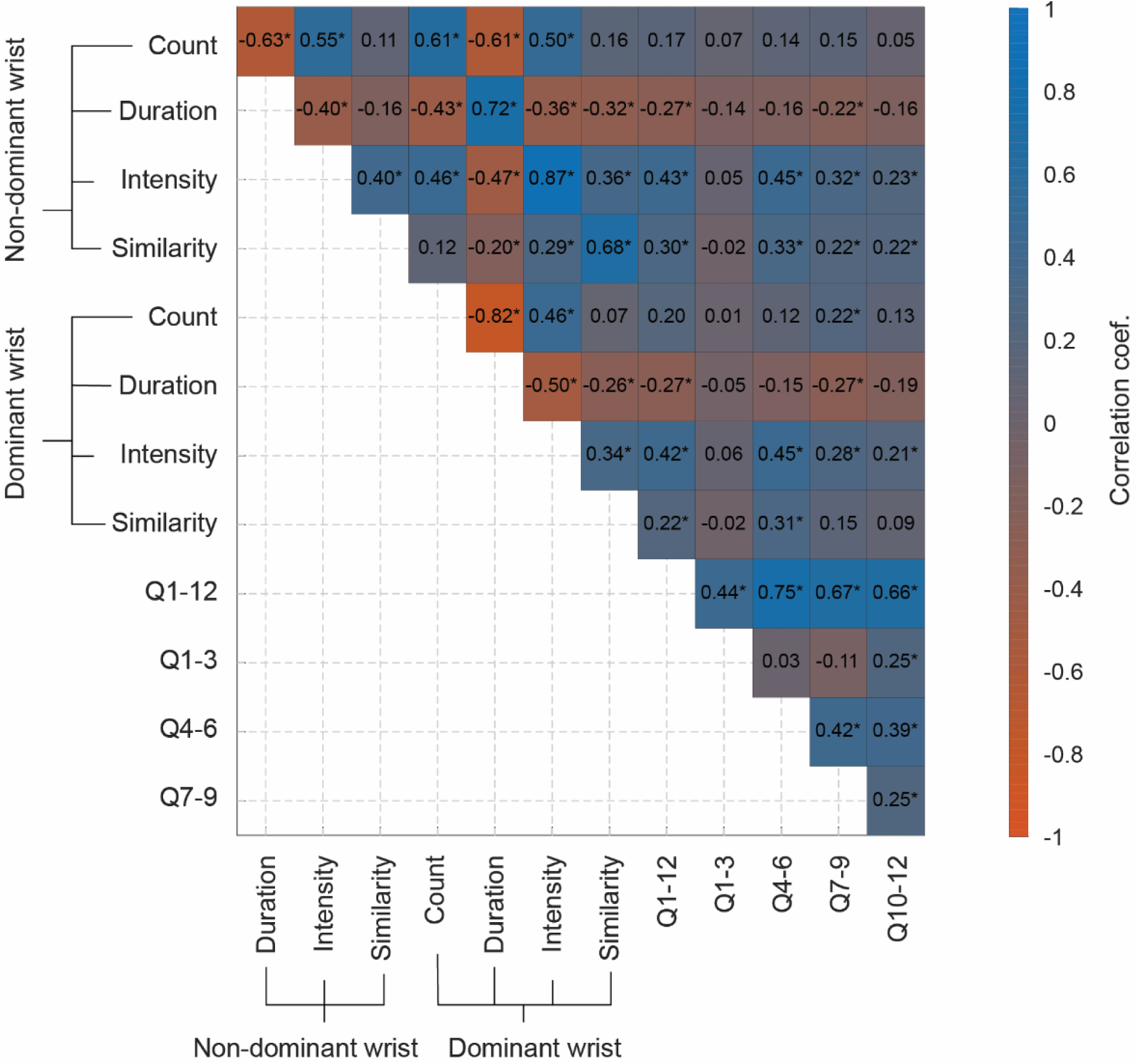
Pearson correlation coefficient between baseline limb swing measures and ALSFRS-RSE total score and subdomain scores (Q1—3 bulbar, Q4—6 fine motor, Q7—9 gross motor, Q10—12 respiratory). ‘*’ indicates statistically significant correlation.

Notable correlations were identified between measures computed from each upper limb. Moderate to strong and negative correlations were identified between count and duration metrics (-0.63 and -0.82 for non-dominant and dominant upper limb, respectively), while moderate negative correlations were observed between intensity and duration metrics (-0.40 and -0.50 for non-dominant and dominant limb, respectively). Moderate positive correlations were observed between count and intensity (0.55 and 0.46 for non-dominant and dominant limb, respectively). These findings indicate that participants who performed more limb swings also executed them more quickly and vigorously, as expected. Strong to very strong positive correlations were observed between metrics from the two limbs (0.61 count, 0.72 duration, 0.87 intensity, 0.68 similarity), indicating similar within-participant upper limb functioning.

### 3.3. Association with time

Using LMMs, we determined that, as expected in this experimental setup, limb swing count remained relatively stable over time in both arms (*p*-value>0.312) (**Table 2**). In contrast, quality metrics – duration, intensity, and similarity – demonstrated statistically significant change over time (all *p*-value<0.001). Specifically, swing duration increased, indicating slower movements, while intensity and similarity measures decreased, highlighting a decline in both vigor and consistency of limb swing repetitions. For both limbs, intensity showed the highest R-squared value, indicating that it was the most accurate and reliable measure for explaining changes in limb function over time.

**Table 2.** Average baseline and weekly change in ALSFRS-RSE scores and the limb swing movements metrics.

| Device location | Outcome | Baseline estimate [95% CI] | Weekly change estimate [95% CI] | *p*-value | R-squared [95% CI] |
| --- | --- | --- | --- | --- | --- |
| Non-dominant wrist |  |  |  |  |  |
|  | Count | 34.66 [33.73, 35.59] | 0.002 [-0.014, 0.019] | 0.773 | 0.758 [0.714, 0.808] |
|  | Duration | 1.408 [1.369, 1.447] | 0.002 [0.001, 0.003] | 5.317e-05 | 0.650 [0.594, 0.702] |
|  | Intensity | 0.6316 [0.5656, 0.6975] | -0.002 [-0.003, -0.001] | 4.515e-06 | 0.882 [0.856, 0.911] |
|  | Similarity | 0.8967 [0.8831, 0.9103] | -0.001 [-0.001, -0.001] | 3.931e-11 | 0.645 [0.607, 0.686] |
| Dominant |  |  |  |  |  |
|  | Count | 34.81 [33.85, 35.78] | 0.008 [-0.008, 0.025] | 0.312 | 0.771 [0.694, 0.820] |
|  | Duration | 1.408 [1.368, 1.448] | 0.001 [0.000, 0.002] | 0.003 | 0.634 [0.575, 0.718] |
|  | Intensity | 0.674 [0.6038, 0.7443] | -0.002 [-0.003, -0.001] | 1.424e-06 | 0.873 [0.842, 0.905] |
|  | Similarity | 0.9076 [0.8957, 0.9195] | -0.001 [-0.001, -0.001] | 3.291e-12 | 0.665 [0.612, 0.726] |
| ALSFRS-RSE |  |  |  |  |  |
|  | Q1—3 | 10.57 [10.34, 10.8] | -0.025 [-0.029, -0.020] | 1.000e-25 | 0.965 [0.953, 0.972] |
|  | Q4—6 | 9.61 [9.345, 9.876] | -0.039 [-0.045, -0.034] | 7.497e-47 | 0.950 [0.942, 0.957] |
|  | Q7—9 | 9.208 [8.932, 9.484] | -0.046 [-0.051, -0.041] | 3.445e-61 | 0.955 [0.947, 0.963] |
|  | Q10—12 | 11.28 [11.11, 11.45] | -0.016 [-0.019, -0.012] | 8.457e-21 | 0.927 [0.907, 0.944] |
|  | Q1—12 | 40.69 [40.09, 41.29] | -0.128 [-0.143, -0.113] | 7.318e-63 | 0.967 [0.958, 0.975] |

A longitudinal evaluation of dominant upper limb swings in one study participant over a 3-year period, using raw accelerometer data collected about every 4 weeks, demonstrated a decline in acceleration and an increase in movement duration over time (**Figure 3**). Of note, this participant also reduced the number of repetitions performed over time, deviating from the group-level trend where swing counts remained relatively stable (upper left panel in **Figure 3c**). Representative patterns of dominant upper limb swing data are presented in the **Supplementary Materials**.

**Figure 3.**
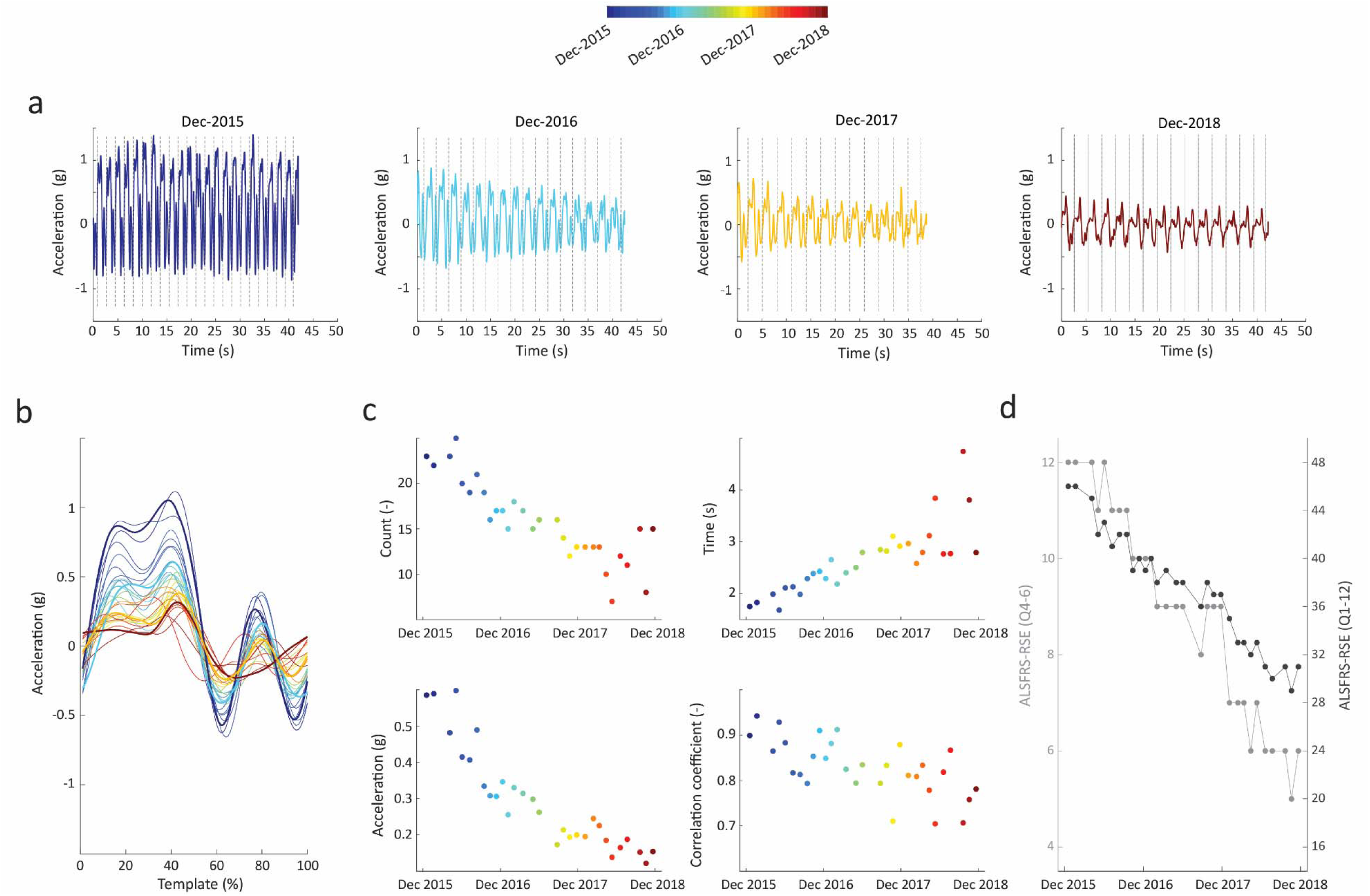
Evaluation of one participant’s dominant upper limb swings over 3 years. a On average, raw accelerometry data (vector magnitude) were collected every 6 weeks. b Data were segmented to obtain individual limb swings templates. Templates were averaged at each time point (all limb swings during exercise, typically 30 or more) to alleviate longitudinal monitoring of disease progression. c We used segmented limb swings to quantify their count, duration, intensity, and similarity. d Upper limb swing metrics were compared with ALSFRS-RSE Q1—12 and Q4—6 responses. Data were derived from the first exercise recorded in December 2015, December 2016, December 2017, and December 2018, i.e., 37 months apart between first and last measurement. During this period, participant decreased their count of upper limb swings (upper left corner of c), intensity (lower left corner of c), and similarity (lower right corner of c), while their duration increased (upper right corner of c) indicating progressing functional decline in upper limb.

### 3.4. Association with ALSFRS-RSE

All limb swing metrics showed significant associations with fine motor function (Table 3), the total ALSFRS-RSE score (Table S1, Supplementary Materials), and the other ALSFRS-RSE subdomains (Tables S2-4, Supplementary Materials). Interestingly, similar results were computed for data collected from both wrists despite survey items such as handwriting (Q4) being specifically tied to dominant-hand activities. Among all, the greatest R-squared was noted from the intensity metrics (0.897 and 0.899 for non-dominant and dominant upper limb, respectively).

**Table 3.** Model intercept and estimated average change associated with a one-point increase in ALSFRS-RSE Q4—6 using limb swing-derived outcomes of accelerometry data for PEs.

| Device location | Movement biomarker | Intercept [95% CI] | Slope [95% CI] | p-value | R-squared [95% CI] |
| --- | --- | --- | --- | --- | --- |
| Non-dominant wrist |  |  |  |  |  |
|  | Count | 7.92 [7.42, 8.42] | 0.020 [0.007, 0.032] | 2.00e-03 | 0.814 [0.769, 0.849] |
|  | Duration | 9.41 [8.92, 9.89] | -0.585 [-0.879, -0.291] | 9.74e-05 | 0.803 [0.766, 0.850] |
|  | Intensity | 7.61 [7.28, 7.94] | 3.73 [3.08, 4.38] | 2.47e-29 | 0.875 [0.848, 0.899] |
|  | Similarity | 5.82 [4.91, 6.73] | 3.08 [2.14, 4.01] | 9.69e-11 | 0.812 [0.745, 0.845] |
| Dominant wrist |  |  |  |  |  |
|  | Count | 7.91 [7.37, 8.44] | 0.019 [0.006, 0.033] | 4.31e-03 | 0.816 [0.772, 0.861] |
|  | Duration | 9.27 [8.77, 9.77] | -0.534 [-0.847, -0.220] | 8.59e-04 | 0.811 [0.769, 0.863] |
|  | Intensity | 7.54 [7.21, 7.87] | 3.76 [3.10, 4.42] | 6.93e-29 | 0.879 [0.860, 0.900] |
|  | Similarity | 5.62 [4.57, 6.68] | 3.27 [2.19, 4.35] | 3.08e-09 | 0.812 [0.768, 0.859] |

### 3.5. Comparison with free-living upper limb movements metrics

The comparative analysis considered data from 193 individuals who recorded at least 21 hours of wear time per day during the free-living monitoring period, representing 41.5% reduction compared to the total sample size and emphasizing challenges associated with adherence related to near-continuous wear. In this subgroup, the baseline total ALSFRS-RSE score was equal to 40.03/48 (SD=5.56), while Q4—6 was equal to 9.44/12 (SD=2.27), similar to the original cohort.

Consistent with previous studies, upper limb movements metrics were significantly associated with responses to the ALSFRS-RSE Q4—6 (Table 4) and Q1—12 scores (Table S5 in Supplementary Materials), regardless of device location on non-dominant or dominant wrist. Interestingly, three out of four (i.e., all except C_fe45_) *p*-values estimated from 21 h of free-living monitoring were greater than for the intensity measure estimated from 45-second exercises. Furthermore, the intensity measure from prescribed upper limbs exercises also demonstrated higher R-squared values, indicating a stronger and more reliable fit to the observed outcomes, compared to free-living movement metrics.

**Table 4.** Model intercept and estimated average change associated with a one-point increase in ALSFRS-RSE Q4—6 using limb swing-derived outcomes of accelerometry data for the free-living upper limb movements. Models were estimated using data from days with simultaneous exercise and substantial sensor wear time (>=21 h).

| Device location | Movement biomarker | Intercept [95% CI] | Slope [95% CI] | p-value | R-squared [95% CI] |
| --- | --- | --- | --- | --- | --- |
| Non-dominant wrist |  |  |  |  |  |
|  | Count | 7.95 [7.29, 8.61] | 0.024 [0.008, 0.041] | 4.27e-03 | 0.796 [0.742, 0.854] |
|  | Duration | 9.79 [9.07, 10.52] | -0.719 [-1.174, -0.265] | 1.94e-03 | 0.806 [0.731, 0.870] |
|  | Intensity | 6.68 [6.13, 7.23] | 3.75 [3.05, 4.45] | 3.20e-25 | 0.899 [0.859, 0.923] |
|  | Similarity | 6.37 [4.92, 7.82] | 2.68 [1.14, 4.23] | 6.12e-04 | 0.798 [0.720, 0.869] |
|  | C<sub>fe45</sub> | 7.03 [6.53, 7.53] | 0.002 [0.001, 0.002] | 4.17e-26 | 0.860 [0.815, 0.907] |
|  | C<sub>sp45</sub> | 7.11 [6.60, 7.63] | 0.001 [0.001, 0.001] | 1.22e-21 | 0.851 [0.800, 0.892] |
|  | D<sub>fe45</sub> | 10.89 [10.39, 11.40] | -3.82 [-4.62, -3.02] | 1.45e-20 | 0.825 [0.763, 0.886] |
|  | D<sub>sp45</sub> | 11.08 [10.53, 11.64] | -6.12 [-7.51, -4.73] | 9.77e-18 | 0.830 [0.765, 0.892] |
| Dominant wrist |  |  |  |  |  |
|  | Count | 8.30 [7.61, 9.00] | 0.010 [-0.007, 0.027] | 0.228 | 0.806 [0.739, 0.861] |
|  | Duration | 9.34 [8.70, 9.98] | -0.469 [-0.852, -0.087] | 0.016 | 0.795 [0.721, 0.857] |
|  | Intensity | 6.72 [6.18, 7.25] | 3.49 [2.86, 4.13] | 7.31e-27 | 0.897 [0.865, 0.921] |
|  | Similarity | 5.68 [4.32, 7.05] | 3.38 [1.96, 4.81] | 3.40e-06 | 0.801 [0.736, 0.870] |
|  | C<sub>fe45</sub> | 6.98 [6.48, 7.49] | 0.002 [0.001, 0.002] | 1.00e-24 | 0.870 [0.825, 0.909] |
|  | C<sub>sp45</sub> | 7.13 [6.62, 7.64] | 0.001 [0.000, 0.001] | 8.84e-18 | 0.855 [0.808, 0.903] |
|  | D<sub>fe45</sub> | 10.58 [10.14, 11.03] | -3.35 [-4.10, -2.61] | 1.29e-18 | 0.864 [0.834, 0.901] |
|  | D<sub>sp45</sub> | 10.85 [10.29, 11.41] | -5.63 [-6.93, -4.32] | 4.01e-17 | 0.848 [0.801, 0.898] |

### 3.6. Impact of limb swing repetitions

Metrics quantified on a limited number of limb swing repetitions demonstrated statistically significant association with the fine motor subdomain (Tables S6-S9 in Supplementary Materials) and the total ALSFRS-RSE score (Tables S10-S13 in Supplementary Materials), even using as few limb swings as five and regardless of device location on non-dominant or dominant wrist. Similar to the above results, the most profound association was determined for the intensity metrics which exhibited in low *p*-value and high narrow coefficient of determination (R-squared).

The three investigated metrics were also significantly associated with time (Tables S14-17 in Supplementary Materials). Unlike the duration and similarity metrics, the intensity metric model showed a very good fit (all R-squared > 0.858) which emphasizes its robustness in estimation of disease status between participants and capacity to track disease status within participants.

## 4. Discussion

In this study, we demonstrated the feasibility of applying brief assessments with DHTs in individuals with ALS, specifically by using wrist-worn accelerometers in home settings to monitor upper limb function during brief PEs. Our analyses covered upper limb function both through PEs and extended free-living periods. Consistent with previous studies showcasing the value of DHTs, including wearable accelerometers and smartphones^12,24,25,38,39^, our findings not only highlight the effectiveness of these technologies but also present a viable approach to minimize participant burden related to extended and continuous wear.

We developed an automated and interpretable signal processing method to analyze data from PEs, based on the time-frequency decomposition via continuous wavelet transformation, estimation of limb swing frequency, and subsequent template matching. We derived four key limb swing movement metrics – count, duration, intensity, and similarity of repetitions – that summarize participants’ exercise performance. We also analyzed four previously developed metrics derived from free-living monitoring using data from at least 21 hours each day^24^. Our findings showed that the prolonged wear-time required for free-living monitoring led to lower adherence and a substantially reduced the valid sample size, indicating that shorter, structured exercise routines may be yield higher than continuous free-living monitoring.

Our results indicate the quality measures of limb swings during PEs, i.e., duration, intensity, and similarity, are significantly associated with longitudinal ALS disease progression as captured by the ALSFRS-RSE. While limb swing counts remained relatively stable, likely due to participants adhering to video instructions, the quality of movements declined over time, reflecting the progressive nature of ALS.

Interestingly, swing intensity showed particularly strong association with survey responses. Compared to free-living metrics, PE limb swing intensity manifested in similarly low *p*-values and higher coefficient of determination (R-squared), suggesting that upper limb function in ALS may be quantified with short, standardized exercises. This approach has the potential to improve study adherence, reduce sample size required in trials, and support clinical management by enabling remote monitoring without the need for all-day wear. This finding highlights the intensity as a potential digital biomarker, capable of capturing clinically meaningful changes in upper limb function. Incorporating intensity into clinical assessments could enable early detection of disease progression and timely interventions.

Our sensitivity analysis indicates that as few as five repetitions during PEs with either limb were sufficient to provide metrics significantly associated with fine motor function and total functional scores. This underscores the value of a wrist-worn accelerometers for capturing brief, yet meaningful, representations of upper limb function.

By reducing the burden of continuous device wear and simplifying data collection protocols, this approach can improve participant adherence and engagement. Enhanced adherence not only ensures higher-quality data but may also decrease the sample size required to achieve statistical power in clinical trials, expediting the evaluation of new therapies.

Several limitations of our study should be acknowledged. First, we lacked a direct method to measure participants’ adherence to the PE protocol in home settings. External factors such as potential inference or assistance from a caregiver may have influenced the recorded measurements. We also had limited control over device placement; despite clear labeling for wrist placement (left wrist, right wrist), participants may have worn devices incorrectly, such as on opposite wrist or even the ankle. To address these limitations, future studies should incorporate real-time monitoring tools or adherence-promoting technologies, such as feedback systems or wearable compliance trackers, to ensure protocol fidelity. Enhanced user training and improved device design could further reduce errors related to improper wear and enhance the accuracy of data collection. Additionally, our analysis focused on a single upper limb movement (shoulder abduction and adduction) which does not fully capture the complexity of compound movements typically used in activities of daily living. Future research should expand to include other upper limb motions, such as elbow flexions and extensions, which are important for functional tasks like for self-feeding.

While this study provides an initial validation for using a wearable device to monitor changes in limb function from brief PEs, establishing clinically meaningful outcomes that resonate with patients and clinicians remains essential^40^. This study was not designed to capture patient perspectives on clinically meaningful changes. Future studies should consider incorporating patient-reported outcome measures (PROMs) or other anchors to enhance the ability of DHT-derived metrics to quantify and predict functional changes meaningful to patients, researchers, and clinicians^41,42^. Lastly, identifying the optimal types, durations, and frequencies of PEs that yield the most predictive value for clinical trials and clinical practice will be crucial, as will considering patient feedback on burden and clinical relevance to ensure adherence^43^.

In conclusion, we demonstrated that short-duration, standardized PE sessions using with wrist-worn accelerometers may be effective tools for quantifying and monitoring upper limb disease progression in ALS. These methods have broad applicability to other conditions characterized by motor decline, offering a flexible and efficient tool for clinical trials and routine care. Combining DHTs with disease-specific quantitative assessments may enhance efficiency in clinical trials and inform clinical decision-making to support improved health outcomes, ultimately contributing to better quality of life for patients.

## 5. Acknowledgments

The authors thank the participants and their caregivers who devoted their time to participate in the study.

## 7. Authors’ contributions

Concept and design – MS, KMB, NC, AP, KTC, FGV, JPO, JDB

Data collection – AP, FGV

Statistical analysis – MS, JPO

Interpretation of results – MS, KMB, NC, KTC, JDB

Manuscript preparation – MS

Critical review of manuscript –KMB, NC, AP, KTC, FGV, JPO, JDB

Study supervision - JDB

## 8. Data availability

Data may be shared upon request and after review and approval by the owners of the data. Shared data will consider deidentified summary statistics used for analytical analysis. Related correspondence should be sent to.

