## Supplementary Tables for "Short prescribed exercises can quantify upper limb functioning in neurodegenerative disease"

### 1. Supplementary Materials

#### 1.1. Association with ALSFRS-RSE

**Table S1**. Model intercept and estimated average change associated with a one-point increase in ALSFRS-RSE Q1—12 using limb swing-derived outcomes of accelerometry data.

| Sensor location | Movement biomarker | Intercept [95% CI] | Slope [95% CI] | *p*-value | R-squared [95% CI] |
| --- | --- | --- | --- | --- | --- |
| Non-dominant wrist |  |  |  |  |  |
|  | Count | 34.80 [33.23, 36.37] | 0.079 [0.039, 0.119] | 1.08e-04 | 0.800 [0.748, 0.841] |
|  | Duration | 40.30 [38.91, 41.69] | -2.066 [-2.994, -1.138] | 1.29e-05 | 0.781 [0.718, 0.856] |
|  | Intensity | 34.34 [33.42, 35.26] | 10.42 [8.71, 12.13] | 1.36e-32 | 0.870 [0.845, 0.904] |
|  | Similarity | 29.66 [26.78, 32.54] | 8.58 [5.66, 11.49] | 7.92e-09 | 0.788 [0.725, 0.861] |
| Dominant wrist |  |  |  |  |  |
|  | Count | 35.13 [33.56, 36.7] | 0.067 [0.027, 0.107] | 1.02e-03 | 0.798 [0.731, 0.849] |
|  | Duration | 39.55 [38.16, 40.93] | -1.624 [-2.565, -0.683] | 7.20e-04 | 0.792 [0.736, 0.842] |
|  | Intensity | 34.30 [33.37, 35.23] | 10.04 [8.32, 11.76] | 4.05e-30 | 0.870 [0.824, 0.903] |
|  | Similarity | 28.60 [25.49, 31.71] | 9.67 [6.49, 12.85] | 2.66e-09 | 0.792 [0.736, 0.842] |

**Table S2**. Model intercept and estimated average change associated with a one-point increase in ALSFRS-RSE Q1—3 using limb swing-derived outcomes of accelerometry data.

| Sensor location | Movement biomarker | Intercept [95% CI] | Slope [95% CI] | *p*-value | R-squared [95% CI] |
| --- | --- | --- | --- | --- | --- |
| Non-dominant wrist |  |  |  |  |  |
|  | Count | 9.41 [8.87, 9.95] | 0.016 [0.004, 0.028] | 9.19e-03 | 0.889 [0.858, 0.916] |
|  | Duration | 10.42 [9.98, 10.85] | -0.350 [-0.627, -0.074] | 0.013 | 0.875 [0.843, 0.918] |
|  | Intensity | 9.34 [8.97, 9.72] | 1.608 [1.208, 2.007] | 3.50e-15 | 0.917 [0.894, 0.936] |
|  | Similarity | 8.74 [7.97, 9.52] | 1.277 [0.542, 2.012] | 6.64e-04 | 0.871 [0.815, 0.913] |
| Dominant wrist |  |  |  |  |  |
|  | Count | 9.49 [9.00, 9.98] | 0.013 [0.002, 0.023] | 0.017 | 0.882 [0.850, 0.918] |
|  | Duration | 10.32 [9.90, 10.74] | -0.302 [-0.561, -0.044] | 0.021 | 0.880 [0.846, 0.909] |
|  | Intensity | 9.37 [9.01, 9.74] | 1.391 [1.042, 1.740] | 6.21e-15 | 0.911 [0.892, 0.932] |
|  | Similarity | 8.31 [7.45, 9.16] | 1.749 [0.922, 2.576] | 3.43e-05 | 0.876 [0.835, 0.917] |

**Table S3**. Model intercept and estimated average change associated with a one-point increase in ALSFRS-RSE Q7—9 using limb swing-derived outcomes of accelerometry data.

| Sensor location | Movement biomarker | Intercept [95% CI] | Slope [95% CI] | *p*-value | R-squared [95% CI] |
| --- | --- | --- | --- | --- | --- |
| Non-dominant wrist |  |  |  |  |  |
|  | Count | 7.117 [6.546, 7.687] | 0.029 [0.014, 0.043] | 9.18e-05 | 0.813 [0.777, 0.859] |
|  | Duration | 9.119 [8.582, 9.657] | -0.751 [-1.107, -0.396] | 3.39e-05 | 0.809 [0.778, 0.842] |
|  | Intensity | 6.956 [6.608, 7.304] | 3.539 [2.935, 4.144] | 2.88e-30 | 0.866 [0.826, 0.889] |
|  | Similarity | 5.159 [4.168, 6.149] | 3.214 [2.228, 4.200] | 1.74e-10 | 0.810 [0.772, 0.858] |
| Dominant wrist |  |  |  |  |  |
|  | Count | 7.276 [6.681, 7.872] | 0.023 [0.008, 0.038] | 2.31e-03 | 0.816 [0.790, 0.866] |
|  | Duration | 8.745 [8.194, 9.296] | -0.514 [-0.877, -0.151] | 5.48e-03 | 0.814 [0.763, 0.851] |
|  | Intensity | 6.97 [6.623, 7.317] | 3.357 [2.745, 3.969] | 8.51e-27 | 0.871 [0.842, 0.893] |
|  | Similarity | 4.977 [3.882, 6.073] | 3.386 [2.274, 4.498] | 2.49e-09 | 0.810 [0.766, 0.850] |

**Table S4**. Model intercept and estimated average change associated with a one-point increase in ALSFRS-RSE Q10—12 using limb swing-derived outcomes of accelerometry data.

| Sensor location | Movement biomarker | Intercept [95% CI] | Slope [95% CI] | *p*-value | R-squared [95% CI] |
| --- | --- | --- | --- | --- | --- |
| Non-dominant wrist |  |  |  |  |  |
|  | Count | 10.34 [9.97, 10.71] | 0.015 [0.006, 0.024] | 7.23e-04 | 0.851 [0.802, 0.885] |
|  | Duration | 11.31 [11.01, 11.62] | -0.340 [-0.528, -0.152] | 3.98e-04 | 0.838 [0.764, 0.886] |
|  | Intensity | 10.43 [10.17, 10.68] | 1.356 [1.010, 1.701] | 1.68e-14 | 0.882 [0.852, 0.912] |
|  | Similarity | 9.715 [9.024, 10.41] | 1.239 [0.558, 1.920] | 3.62e-04 | 0.845 [0.796, 0.889] |
| Dominant wrist |  |  |  |  |  |
|  | Count | 10.46 [10.08, 10.84] | 0.012 [0.002, 0.021] | 0.013 | 0.853 [0.805, 0.896] |
|  | Duration | 11.2 [10.91, 11.49] | -0.266 [-0.446, -0.085] | 3.91e-03 | 0.846 [0.787, 0.885] |
|  | Intensity | 10.43 [10.17, 10.69] | 1.305 [0.952, 1.658] | 4.75e-13 | 0.882 [0.828, 0.911] |
|  | Similarity | 9.578 [8.945, 10.21] | 1.384 [0.766, 2.001] | 1.13e-05 | 0.849 [0.778, 0.896] |

#### 1.2. Exercise vs. free-living upper limb movements metrics

**Table S5**. Model intercept and estimated average change associated with a one-point increase in ALSFRS-RSE Q1—12 using limb swing-derived outcomes of accelerometry data for the free-living upper limb movements. Models were estimated using data from days with simultaneous exercise and substantial sensor wear time (>=21 h).

| Sensor location | Movement biomarker | Intercept [95% CI] | Slope [95% CI] | *p*-value | R-squared [95% CI] |
| --- | --- | --- | --- | --- | --- |
| Non-dominant wrist |  |  |  |  |  |
|  | Count | 35.54 [33.42, 37.65] | 0.070 [0.017, 0.123] | 9.51e-03 | 0.811 [0.707, 0.889] |
|  | Duration | 40.62 [38.36, 42.88] | -1.910 [-3.473, -0.346] | 0.017 | 0.826 [0.739, 0.891] |
|  | Intensity | 32.10 [30.36, 33.84] | 10.33 [8.15, 12.52] | 3.12e-20 | 0.908 [0.861, 0.943] |
|  | Similarity | 31.73 [27.26, 36.19] | 6.74 [2.07, 11.40] | 4.65e-03 | 0.792 [0.639, 0.890] |
|  | C_fe45_ | 32.71 [31.22, 34.20] | 0.004 [0.004, 0.005] | 1.47e-24 | 0.877 [0.819, 0.920] |
|  | C_sp45_ | 32.78 [31.22, 34.33] | 0.002 [0.002, 0.002] | 6.36e-22 | 0.869 [0.810, 0.915] |
|  | D_fe45_ | 43.80 [42.44, 45.15] | -11.02 [-13.26, -8.77] | 1.24e-21 | 0.821 [0.713, 0.912] |
|  | D_sp45_ | 44.27 [42.66, 45.87] | -17.27 [-21.64, -12.91] | 1.14e-14 | 0.834 [0.723, 0.915] |
| Dominant wrist |  |  |  |  |  |
|  | Count | 36.40 [34.30, 38.51] | 0.037 [-0.016, 0.090] | 0.1687 | 0.813 [0.729, 0.902] |
|  | Duration | 39.72 [37.82, 41.62] | -1.415 [-2.644, -0.187] | 0.023 | 0.805 [0.661, 0.885] |
|  | Intensity | 32.26 [30.61, 33.90] | 9.682 [7.742, 11.623] | 2.93e-22 | 0.899 [0.857, 0.936] |
|  | Similarity | 29.23 [25.38, 33.07] | 9.456 [5.476, 13.436] | 3.33e-06 | 0.791 [0.616, 0.885] |
|  | C_fe45_ | 32.85 [31.40, 34.29] | 0.004 [0.004, 0.005] | 8.35e-25 | 0.873 [0.797, 0.923] |
|  | C_sp45_ | 32.98 [31.44, 34.52] | 0.002 [0.001, 0.002] | 2.37e-19 | 0.866 [0.810, 0.914] |
|  | D_fe45_ | 43.25 [42.10, 44.41] | -10.10 [-12.26, -7.94] | 9.75e-20 | 0.865 [0.790, 0.909] |
|  | D_sp45_ | 43.78 [42.23, 45.34] | -15.94 [-20.07, -11.82] | 4.65e-14 | 0.847 [0.771, 0.913] |

#### 1.3. Association based on fewer limb swings

##### 1.3.1. Association with ALSFRS-RSE Q4—6

**Table S6.** Model intercept and estimated average change associated with a one-point increase in ALSFRS-RSE Q4—6 using accelerometry outcomes quantified from 5 initial limb swings.

| Sensor location | Movement biomarker | Intercept [95% CI] | Slope [95% CI] | *p*-value | R-squared [95% CI] |
| --- | --- | --- | --- | --- | --- |
| Non-dominant wrist |  |  |  |  |  |
|  | Duration | 8.61 [8.33, 8.88] | -0.035 [-0.048, -0.021] | 7.23e-07 | 0.774 [0.728, 0.836] |
|  | Intensity | 7.77 [7.46, 8.09] | 3.107 [2.516, 3.699] | 9.16e-25 | 0.867 [0.835, 0.889] |
|  | Similarity | 7.46 [6.87, 8.04] | 1.234 [0.712, 1.755] | 3.58e-06 | 0.791 [0.732, 0.834] |
| Dominant wrist |  |  |  |  |  |
|  | Duration | 8.61 [8.34, 8.88] | -0.038 [-0.059, -0.018] | 2.77e-05 | 0.772 [0.735, 0.826] |
|  | Intensity | 7.76 [7.43, 8.08] | 2.975 [2.414, 3.536] | 3.24e-25 | 0.866 [0.839, 0.890] |
|  | Similarity | 7.89 [7.34, 8.46] | 0.748 [0.228, 1.268] | 4.84e-03 | 0.787 [0.744, 0.853] |

**Table S7.** Model intercept and estimated average change associated with a one-point increase in ALSFRS-RSE Q4—6 using accelerometry outcomes quantified from 10 initial limb swings.

| Sensor location | Movement biomarker | Intercept [95% CI] | Slope [95% CI] | *p*-value | R-squared [95% CI] |
| --- | --- | --- | --- | --- | --- |
| Non-dominant wrist |  |  |  |  |  |
|  | Duration | 8.60 [8.36, 8.88] | -0.031 [-0.044, -0.018] | 2.82e-06 | 0.772 [0.729, 0.847] |
|  | Intensity | 7.78 [7.46, 8.09] | 3.180 [2.582, 3.778] | 2.54e-25 | 0.869 [0.847, 0.895] |
|  | Similarity | 7.42 [6.66, 8.18] | 1.315 [0.575, 2.054] | 4.96e-04 | 0.798 [0.765, 0.840] |
| Dominant wrist |  |  |  |  |  |
|  | Duration | 8.60 [8.33, 8.87] | -0.031 [-0.050, -0.012] | 1.64e-03 | 0.772 [0.715, 0.827] |
|  | Intensity | 7.74 [7.42, 8.06] | 3.069 [2.492, 3.645] | 2.06e-25 | 0.869 [0.840, 0.899] |
|  | Similarity | 7.36 [6.64, 8.08] | 1.359 [0.653, 2.066] | 1.63e-04 | 0.795 [0.749, 0.838] |

**Table S8.** Model intercept and estimated average change associated with a one-point increase in ALSFRS-RSE Q4—6 using accelerometry outcomes quantified from 15 initial limb swings.

| Sensor location | Movement biomarker | Intercept [95% CI] | Slope [95% CI] | *p*-value | R-squared [95% CI] |
| --- | --- | --- | --- | --- | --- |
| Non-dominant wrist |  |  |  |  |  |
|  | Duration | 8.65 [8.25, 9.06] | -0.041 [-0.252, 0.170] | 0.701 | 0.792 [0.740, 0.833] |
|  | Intensity | 7.79 [7.47, 8.11] | 3.230 [2.618, 3.842] | 5.78e-25 | 0.873 [0.840, 0.896] |
|  | Similarity | 7.11 [6.25, 7.97] | 1.677 [0.826, 2.528] | 1.13e-04 | 0.804 [0.762, 0.851] |
| Dominant wrist |  |  |  |  |  |
|  | Duration | 8.63 [8.35, 8.90] | -0.035 [-0.059, -0.011] | 4.36e-03 | 0.778 [0.753, 0.834] |
|  | Intensity | 7.75 [7.43, 8.07] | 3.086 [2.518, 3.653] | 1.99e-26 | 0.874 [0.853, 0.897] |
|  | Similarity | 6.89 [6.02, 7.76] | 1.889 [1.012, 2.766] | 2.43e-05 | 0.804 [0.750, 0.841] |

**Table S9.** Model intercept and estimated average change associated with a one-point increase in ALSFRS-RSE Q4—6 using accelerometry outcomes quantified from 20 initial limb swings.

| Sensor location | Movement biomarker | Intercept [95% CI] | Slope [95% CI] | *p*-value | R-squared [95% CI] |
| --- | --- | --- | --- | --- | --- |
| Non-dominant wrist |  |  |  |  |  |
|  | Duration | 8.68 [8.19, 9.17] | -0.056 [-0.341, 0.229] | 0.700 | 0.803 [0.744, 0.851] |
|  | Intensity | 7.83 [7.51, 8.15] | 2.981 [2.434, 3.528] | 1.64e-26 | 0.873 [0.843, 0.898] |
|  | Similarity | 6.79 [5.84, 7.73] | 2.070 [1.117, 3.022] | 2.06e-05 | 0.811 [0.755, 0.855] |
| Dominant wrist |  |  |  |  |  |
|  | Duration | 8.48 [8.07, 8.89] | 0.081 [-0.127, 0.288] | 0.447 | 0.794 [0.731, 0.837] |
|  | Intensity | 7.76 [7.44, 8.08] | 2.906 [2.396, 3.416] | 8.07e-29 | 0.874 [0.845, 0.898] |
|  | Similarity | 6.70 [5.81, 7.59] | 2.112 [1.206, 3.018] | 4.89e-06 | 0.808 [0.769, 0.854] |

##### 2.1.1. Association with ALSFRS-RSE Q1—12

**Table S10.** Model intercept and estimated average change associated with a one-point increase in ALSFRS-RSE Q1—12 using accelerometry outcomes quantified from 5 initial limb swings.

| Sensor location | Movement biomarker | Intercept [95% CI] | Slope [95% CI] | *p*-value | R-squared [95% CI] |
| --- | --- | --- | --- | --- | --- |
| Non-dominant wrist |  |  |  |  |  |
|  | Duration | 37.47 [36.79, 38.14] | -0.106 [-0.154, -0.058] | 1.48e-05 | 0.741 [0.685, 0.813] |
|  | Intensity | 34.89 [34.01, 35.78] | 8.691 [7.104, 10.278] | 9.09e-27 | 0.857 [0.812, 0.887] |
|  | Similarity | 34.29 [32.53, 36.05] | 3.375 [1.742, 5.007] | 5.10e-05 | 0.759 [0.696, 0.813] |
| Dominant wrist |  |  |  |  |  |
|  | Duration | 37.50 [36.84, 38.17] | -0.144 [-0.220, -0.067] | 2.20e-04 | 0.741 [0.671, 0.806] |
|  | Intensity | 34.92 [34.04, 35.81] | 8.189 [6.641, 9.738] | 4.43e-25 | 0.853 [0.818, 0.890] |
|  | Similarity | 35.56 [33.87, 37.26] | 1.962 [0.343, 3.581] | 0.017 | 0.758 [0.669, 0.815] |

**Table S11.** Model intercept and estimated average change associated with a one-point increase in ALSFRS-RSE Q1—12 using accelerometry outcomes quantified from 10 initial limb swings.

| Sensor location | Movement biomarker | Intercept [95% CI] | Slope [95% CI] | *p*-value | R-squared [95% CI] |
| --- | --- | --- | --- | --- | --- |
| Non-dominant wrist |  |  |  |  |  |
|  | Duration | 37.46 [36.79, 38.13] | -0.095 [-0.142, -0.048] | 8.31e-05 | 0.734 [0.693, 0.819] |
|  | Intensity | 34.90 [34.02, 35.78] | 8.765 [7.198, 10.333] | 7.85e-28 | 0.857 [0.811, 0.889] |
|  | Similarity | 34.26 [31.96, 36.57] | 3.545 [1.262, 5.829] | 2.34e-03 | 0.764 [0.672, 0.820] |
| Dominant wrist |  |  |  |  |  |
|  | Duration | 37.46 [36.80, 38.13] | -0.110 [-0.175, -0.046] | 7.53e-04 | 0.731 [0.692, 0.799] |
|  | Intensity | 34.88 [34, 35.76] | 8.419 [6.852, 9.986] | 7.75e-26 | 0.854 [0.832, 0.887] |
|  | Similarity | 33.96 [31.72, 36.21] | 3.776 [1.549, 6.003] | 8.89e-04 | 0.763 [0.679, 0.826] |

**Table S12.** Model intercept and estimated average change associated with a one-point increase in ALSFRS-RSE Q1—12 using accelerometry outcomes quantified from 15 initial limb swings.

| Sensor location | Movement biomarker | Intercept [95% CI] | Slope [95% CI] | *p*-value | R-squared [95% CI] |
| --- | --- | --- | --- | --- | --- |
| Non-dominant wrist |  |  |  |  |  |
|  | Duration | 37.92 [36.78, 39.06] | -0.368 [-1.049, 0.313] | 0.289 | 0.747 [0.684, 0.822] |
|  | Intensity | 34.94 [34.06, 35.82] | 8.780 [7.228, 10.333] | 1.97e-28 | 0.856 [0.826, 0.888] |
|  | Similarity | 33.24 [30.77, 35.71] | 4.713 [2.260, 7.165] | 1.66e-04 | 0.762 [0.697, 0.820] |
| Dominant wrist |  |  |  |  |  |
|  | Duration | 37.54 [36.87, 38.20] | -0.122 [-0.196, -0.047] | 1.42e-03 | 0.728 [0.682, 0.805] |
|  | Intensity | 34.91 [34.03, 35.78] | 8.284 [6.802, 9.767] | 8.48e-28 | 0.852 [0.819, 0.885] |
|  | Similarity | 32.50 [29.92, 35.08] | 5.425 [2.827, 8.024] | 4.30e-05 | 0.764 [0.706, 0.815] |

**Table S13.** Model intercept and estimated average change associated with a one-point increase in ALSFRS-RSE Q1—12 using accelerometry outcomes quantified from 20 initial limb swings.

| Sensor location | Movement biomarker | Intercept [95% CI] | Slope [95% CI] | *p*-value | R-squared [95% CI] |
| --- | --- | --- | --- | --- | --- |
| Non-dominant wrist |  |  |  |  |  |
|  | Duration | 38.21 [36.84, 39.58] | -0.539 [-1.433, 0.355] | 0.237 | 0.746 [0.687, 0.810] |
|  | Intensity | 35.07 [34.19, 35.94] | 8.186 [6.770, 9.603] | 1.32e-29 | 0.847 [0.814, 0.878] |
|  | Similarity | 32.47 [29.67, 35.28] | 5.686 [2.867, 8.506] | 7.75e-05 | 0.755 [0.680, 0.809] |
| Dominant wrist |  |  |  |  |  |
|  | Duration | 37.36 [36.27, 38.45] | 0.036 [-0.573, 0.646] | 0.906 | 0.736 [0.701, 0.801] |
|  | Intensity | 34.95 [34.07, 35.83] | 7.909 [6.544, 9.273] | 9.23e-30 | 0.847 [0.817, 0.879] |
|  | Similarity | 31.95 [29.35, 34.55] | 6.102 [3.451, 8.753] | 6.49e-06 | 0.758 [0.698, 0.816] |

##### 2.1.2. Association with time

**Table S14.** Average baseline and weekly change in ALSFRS-RSE scores and accelerometry outcomes quantified from 5 initial limb swing movements.

| Sensor location | Outcome | Baseline estimate [95% CI] | Weekly change estimate [95% CI] | *p*-value | R-squared [95% CI] |
| --- | --- | --- | --- | --- | --- |
| Non-dominant wrist |  |  |  |  |  |
|  | Duration | 1.561 [1.432, 1.690] | 0.009 [0.005, 0.014] | 1.89e-05 | 0.201 [0.185, 0.254] |
|  | Intensity | 0.685 [0.614, 0.757] | -0.002 [-0.003, -0.001] | 7.40e-04 | 0.870 [0.851, 0.894] |
|  | Similarity | 0.921 [0.814, 1.028] | 0.001 [-0.001, 0.004] | 0.341 | 0.154 [0.084, 0.244] |
| Dominant wrist |  |  |  |  |  |
|  | Duration | 1.572 [1.455, 1.688] | 0.008 [0.004, 0.011] | 2.14e-05 | 0.178 [0.121, 0.252] |
|  | Intensity | 0.721 [0.646, 0.796] | -0.002 [-0.003, -0.001] | 1.39e-04 | 0.858 [0.832, 0.888] |
|  | Similarity | 0.906 [0.896, 0.916] | -0.001 [-0.001, -0.000] | 7.45e-06 | 0.424 [0.403, 0.515] |

**Table S15.** Average baseline and weekly change in ALSFRS-RSE scores and accelerometry outcomes quantified from 10 initial limb swing movements.

| Sensor location | Outcome | Baseline estimate [95% CI] | Weekly change estimate [95% CI] | *p*-value | R-squared [95% CI] |
| --- | --- | --- | --- | --- | --- |
| Non-dominant wrist |  |  |  |  |  |
|  | Duration | 1.564 [1.451, 1.677] | 0.007 [0.003, 0.011] | 1.48e-04 | 0.179 [0.135, 0.236] |
|  | Intensity | 0.680 [0.609, 0.751] | -0.002 [-0.003, -0.001] | 1.47e-04 | 0.872 [0.851, 0.893] |
|  | Similarity | 0.906 [0.894, 0.917] | -0.001 [-0.001, -0.000] | 1.62e-06 | 0.573 [0.525, 0.627] |
| Dominant wrist |  |  |  |  |  |
|  | Duration | 1.595 [1.483, 1.706] | 0.005 [0.002, 0.008] | 3.54e-04 | 0.113 [0.086, 0.173] |
|  | Intensity | 0.719 [0.643, 0.794] | -0.002 [-0.003, -0.001] | 3.94e-05 | 0.863 [0.827, 0.887] |
|  | Similarity | 0.911 [0.900, 0.921] | -0.001 [-0.001, -0.000] | 8.24e-08 | 0.590 [0.525, 0.659] |

**Table S16.** Average baseline and weekly change in ALSFRS-RSE scores and accelerometry outcomes quantified from 15 initial limb swing movements.

| Sensor location | Outcome | Baseline estimate [95% CI] | Weekly change estimate [95% CI] | *p*-value | R-squared [95% CI] |
| --- | --- | --- | --- | --- | --- |
| Non-dominant wrist |  |  |  |  |  |
|  | Duration | 1.504 [1.397, 1.611] | 0.006 [0.003, 0.010] | 1.03e-03 | 0.136 [0.130, 0.202] |
|  | Intensity | 0.673 [0.602, 0.743] | -0.002 [-0.003, -0.001] | 4.52e-05 | 0.871 [0.848, 0.895] |
|  | Similarity | 0.907 [0.895, 0.915] | -0.001 [-0.001, -0.001] | 4.52e-08 | 0.610 [0.569, 0.661] |
| Dominant wrist |  |  |  |  |  |
|  | Duration | 1.526 [1.436, 1.615] | 0.004 [0.001, 0.006] | 1.68e-03 | 0.061 [0.056, 0.144] |
|  | Intensity | 0.712 [0.637, 0.788] | -0.002 [-0.003, -0.001] | 1.54e-05 | 0.863 [0.832, 0.891] |
|  | Similarity | 0.913 [0.901, 0.924] | -0.001 [-0.001, -0.001] | 1.87e-09 | 0.631 [0.582, 0.690] |

**Table S17.** Average baseline and weekly change in ALSFRS-RSE scores and accelerometry outcomes quantified from 20 initial limb swing movements.

| Sensor location | Outcome | Baseline estimate [95% CI] | Weekly change estimate [95% CI] | *p*-value | R-squared [95% CI] |
| --- | --- | --- | --- | --- | --- |
| Non-dominant wrist |  |  |  |  |  |
|  | Duration | 1.389 [1.298, 1.480] | 0.006 [0.002, 0.010] | 5.98e-03 | 0.139 [0.117, 0.217] |
|  | Intensity | 0.667 [0.597, 0.737] | -0.002 [-0.003, -0.001] | 3.27e-05 | 0.869 [0.851, 0.900] |
|  | Similarity | 0.902 [0.889, 0.916] | -0.001 [-0.001, -0.001] | 7.77e-09 | 0.644 [0.600, 0.694] |
| Dominant wrist |  |  |  |  |  |
|  | Duration | 1.426 [1.340, 1.511] | 0.003 [-0.001, 0.006] | 0.102 | 0.093 [0.088, 0.247] |
|  | Intensity | 0.709 [0.635, 0.784] | -0.002 [-0.003, -0.001] | 9.41e-06 | 0.862 [0.837, 0.884] |
|  | Similarity | 0.908 [0.896, 0.921] | -0.001 [-0.001, -0.001] | 6.13e-10 | 0.665 [0.612, 0.727] |
