## Supplementary figures and images for "Short prescribed exercises can quantify upper limb functioning in neurodegenerative disease"

### Supplementary Figures

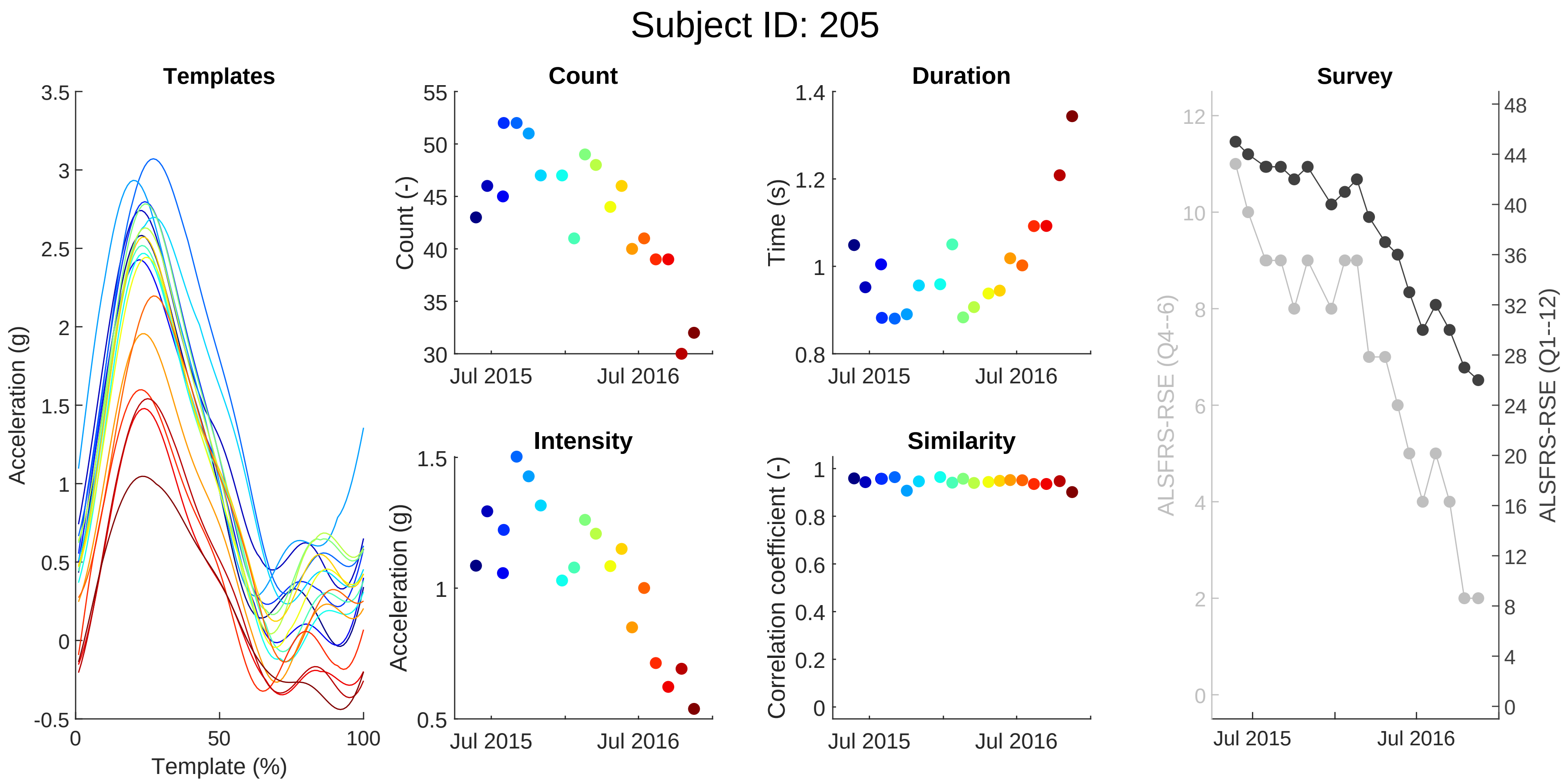

#### Subject ID: 236

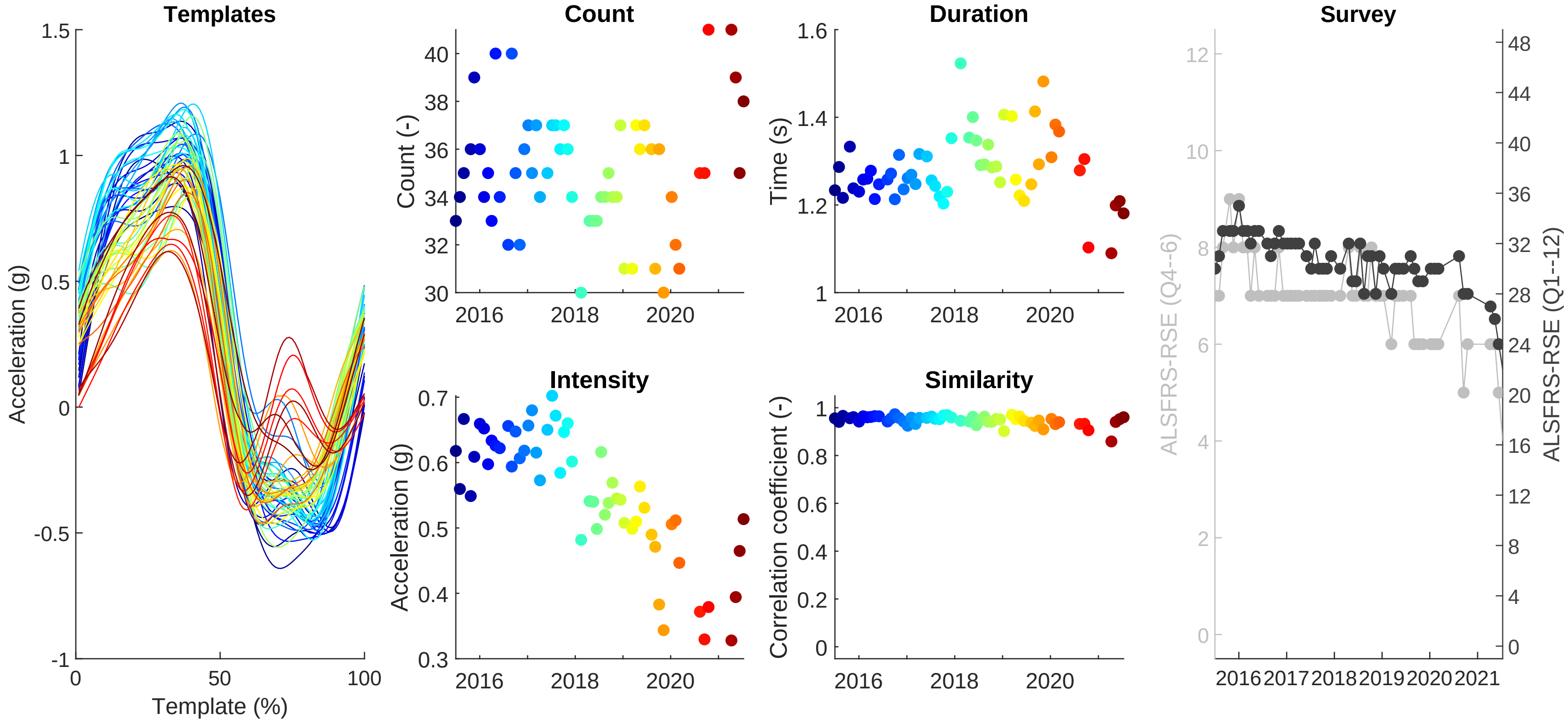

Subject ID: 319

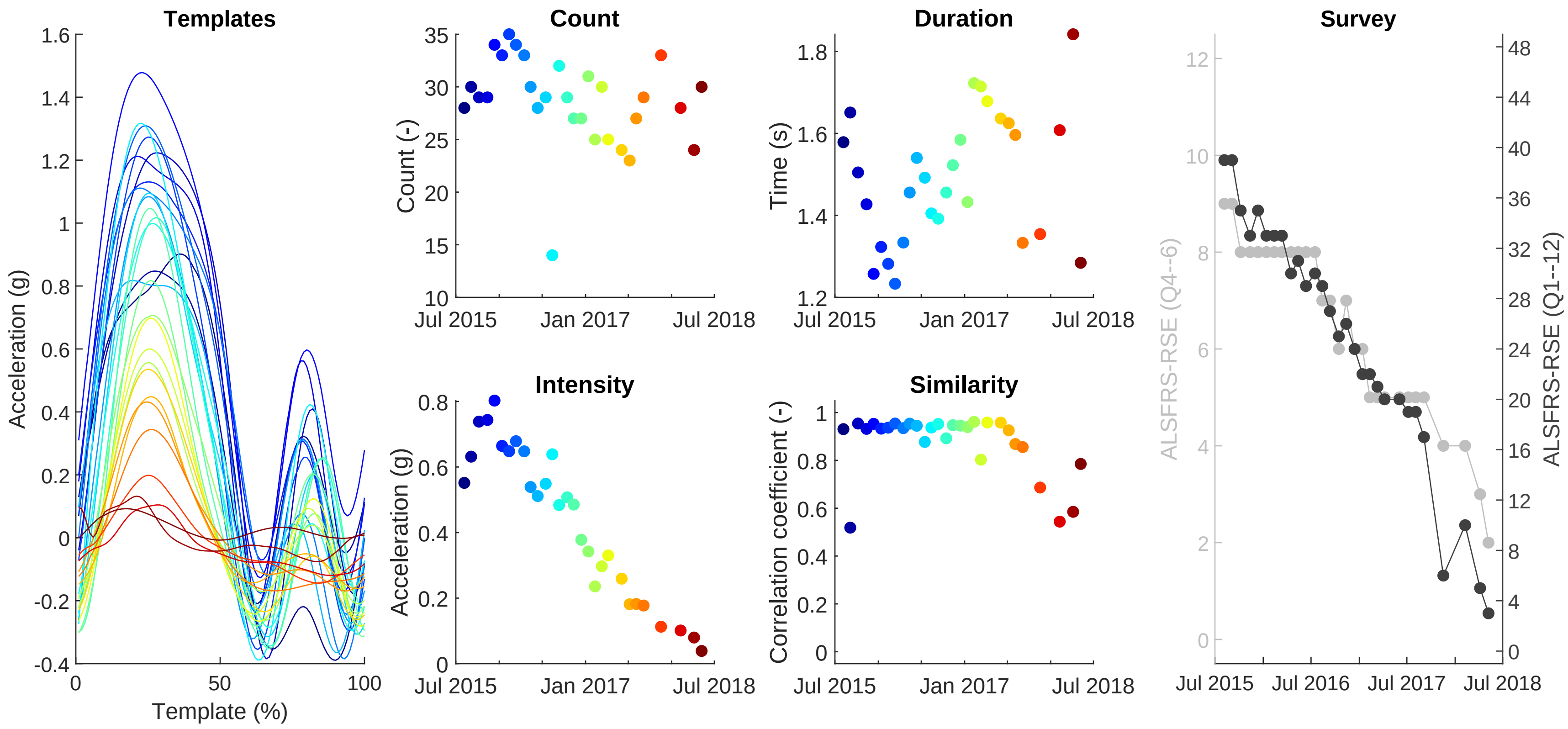

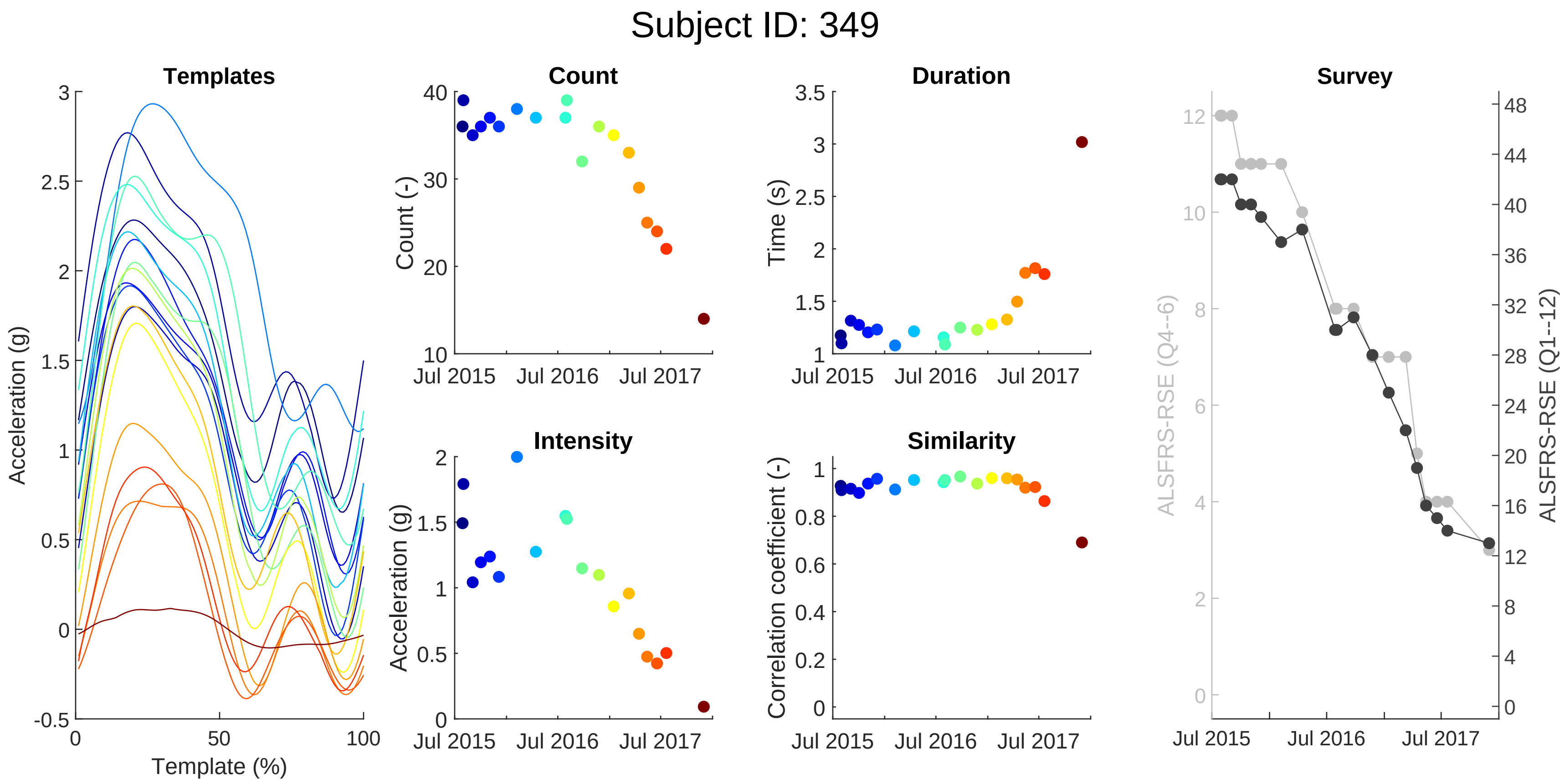

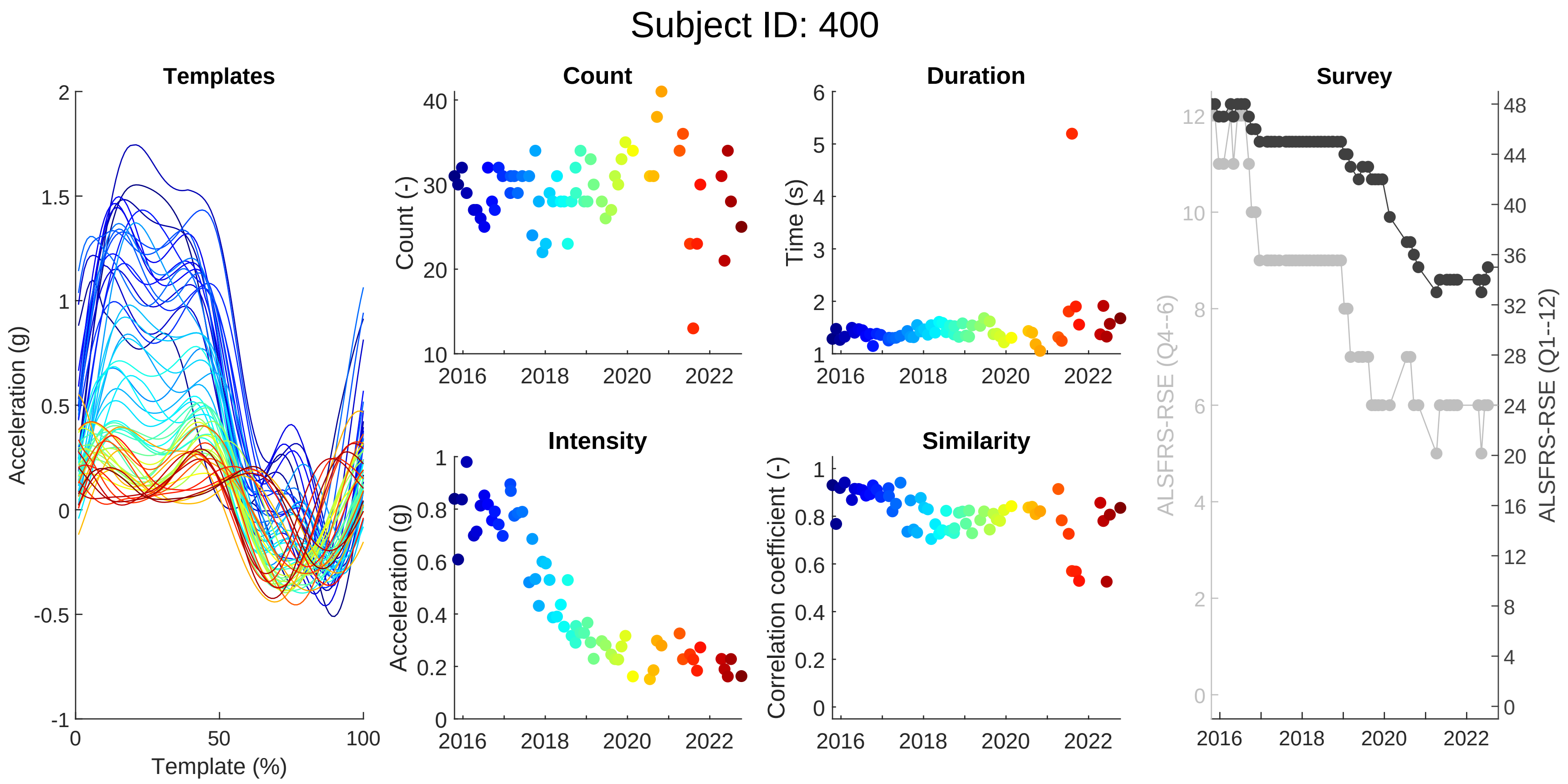

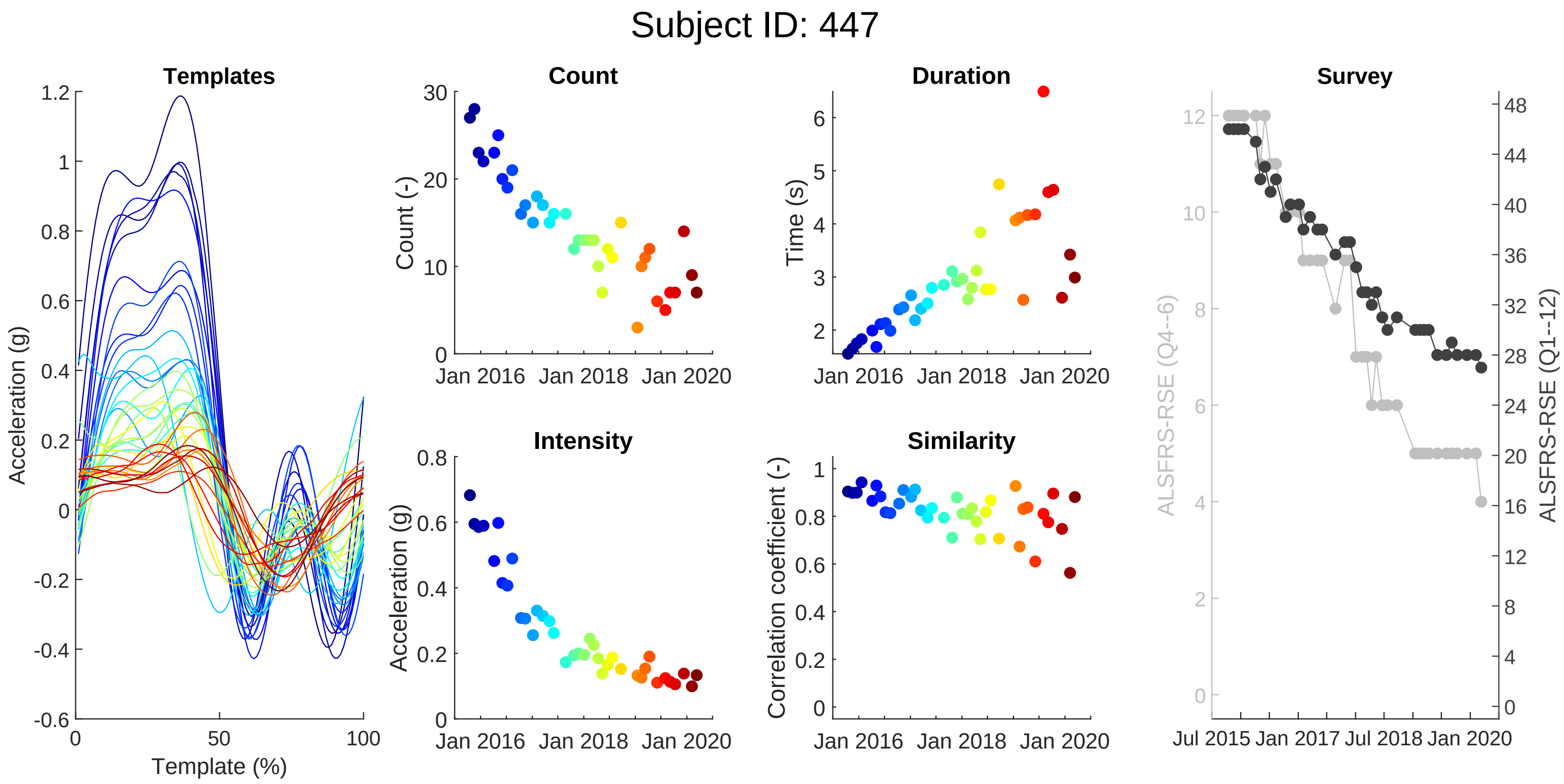

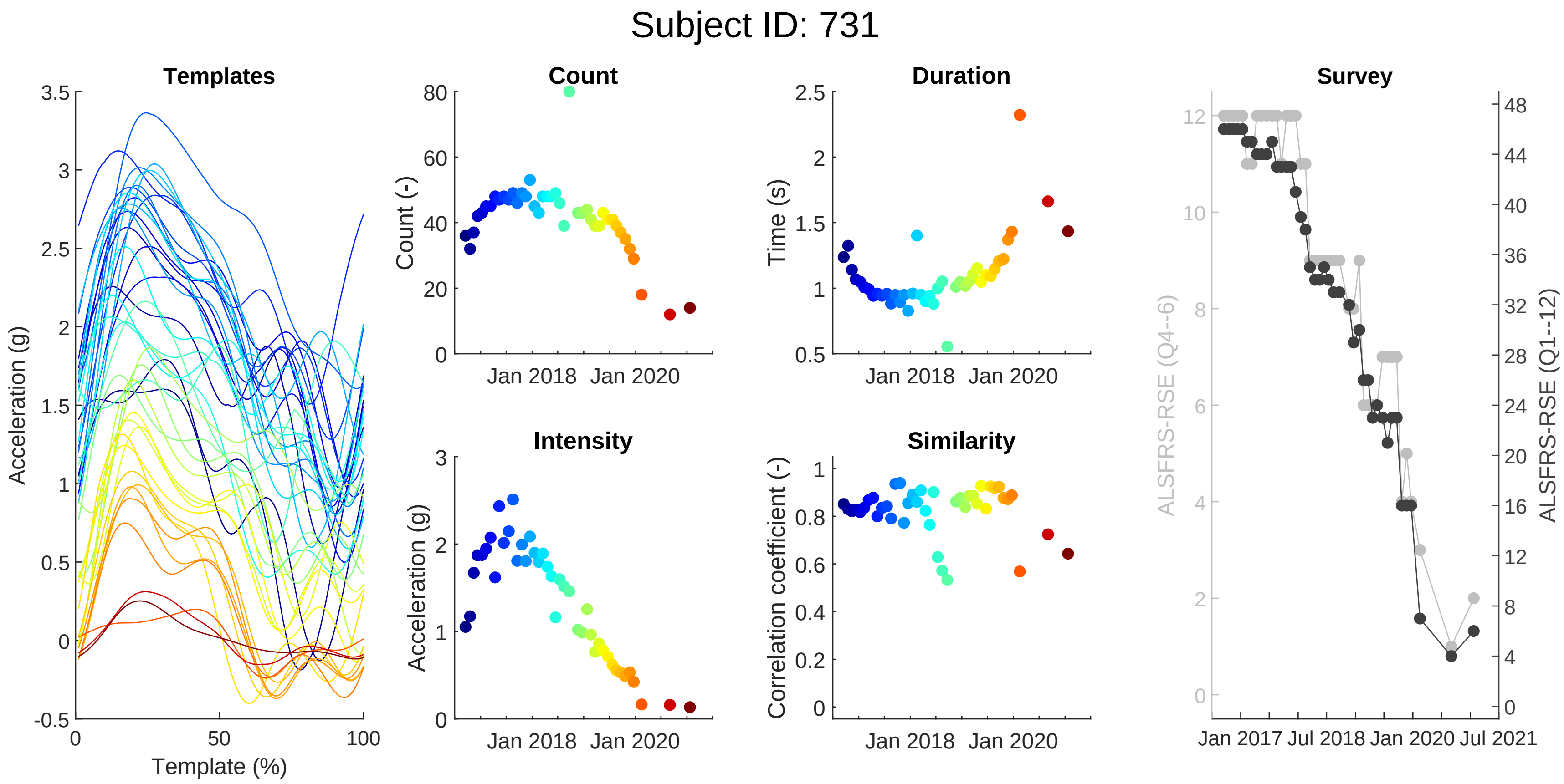

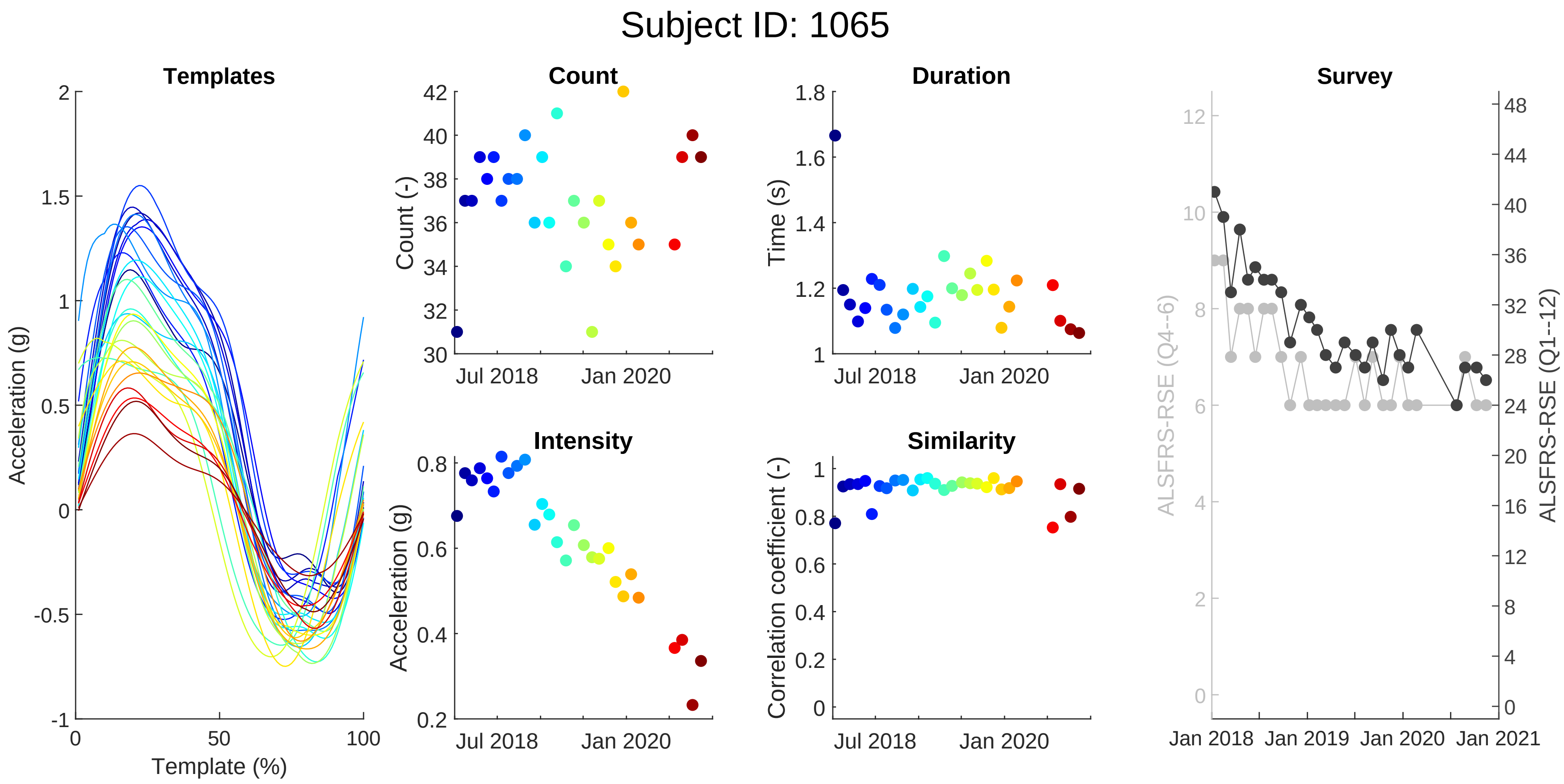

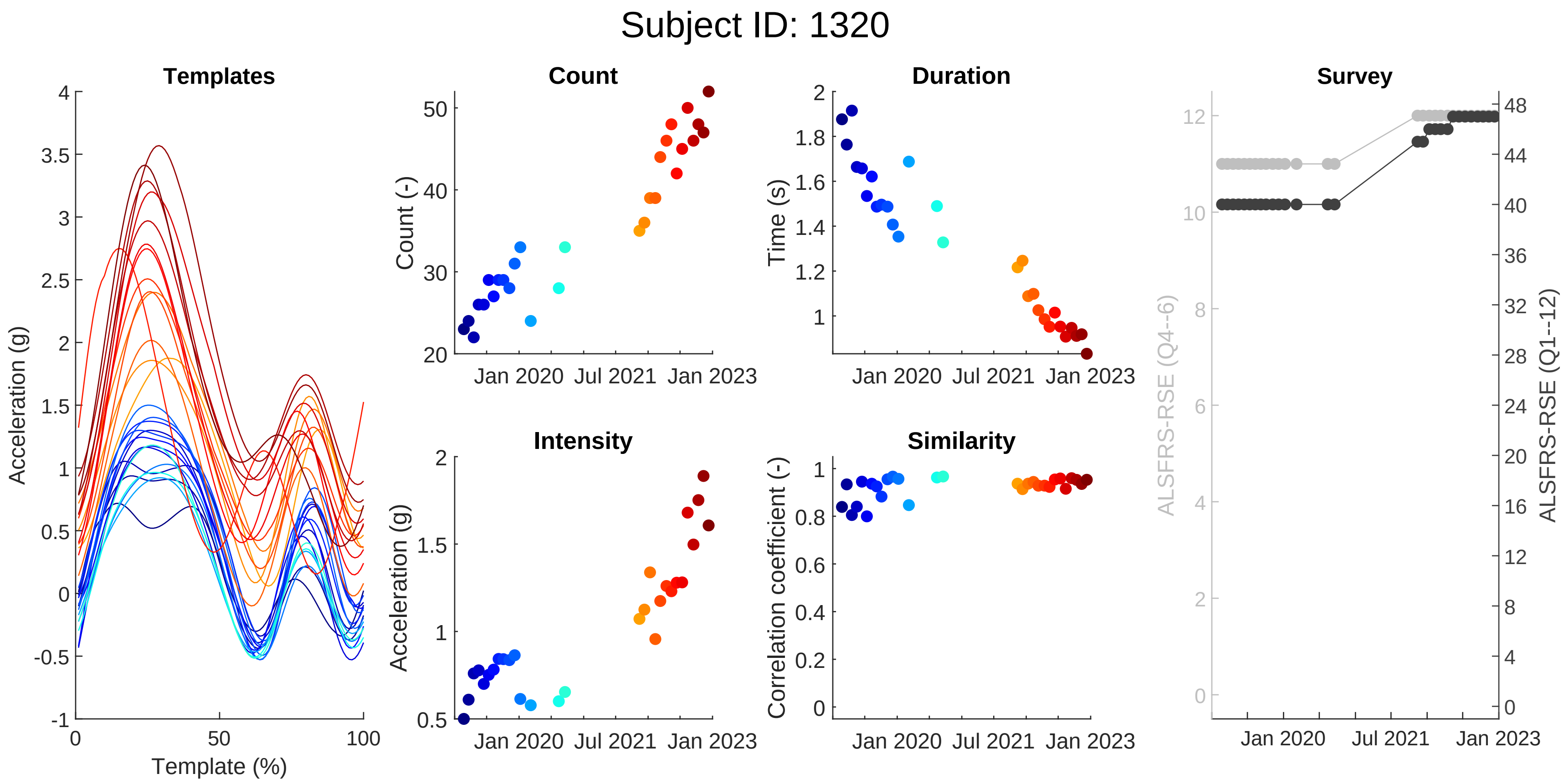

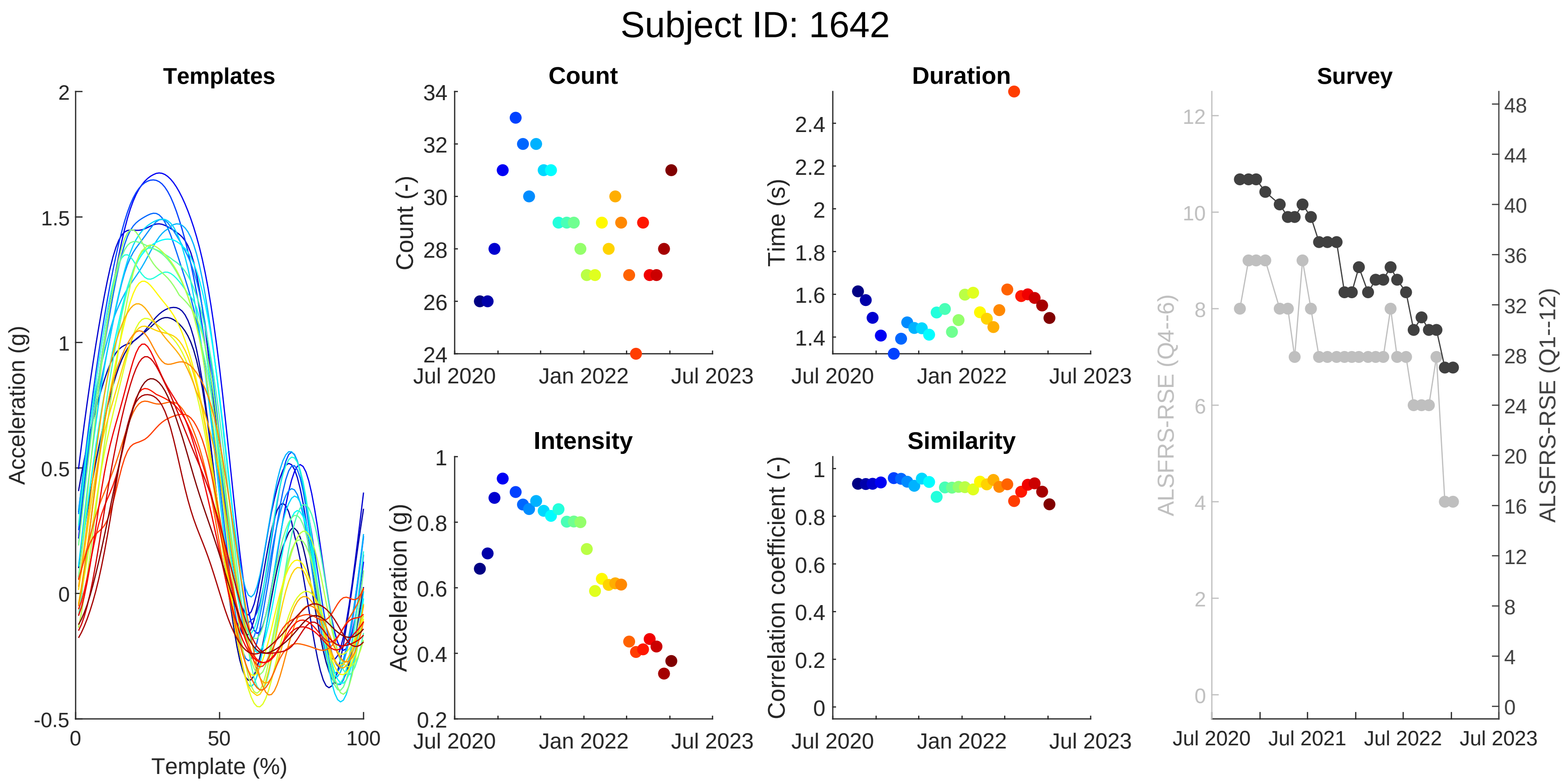
